# Neurodevelopmental disorders associated variants in *ADAT3* disrupt the activity of the ADAT2/ADAT3 tRNA deaminase complex and impair neuronal migration

**DOI:** 10.1101/2024.03.01.24303485

**Authors:** Jordi Del-Pozo-Rodriguez, Peggy Tilly, Romain Lecat, Hugo Rolando Vaca, Laureline Mosser, Elena Brivio, Till Balla, Marina Vitoria Gomes, Elizabeth Ramos-Morales, Noémie Schwaller, Thalia Salinas-Giegé, Grace VanNoy, Eleina M. England, Alysia Kern Lovgren, Melanie O’Leary, Maya Chopra, Naomi Meave Ojeda, Mehran Beiraghi Toosi, Atieh Eslahi, Masoome Alerasool, Majid Mojarrad, Lynn S. Pais, Rebecca C. Yeh, Dustin L. Gable, Mais O Hashem, Firdous Abdulwahab, Hamad Alzaidan, Hesham Aldhalaan, Ehab Tous, Afaf Alsagheir, Mohammed Alowain, Abdullah Tamim, Khowlah Alfayez, Amal Alhashem, Aisha Alnuzha, Mona Kamel, Bashayer S Al-Awam, Walaa Elnaggar, Nihal Almenabawy, Anne O’Donnell-Luria, Jennifer E. Neil, Joseph G. Gleeson, Christopher A. Walsh, Fowzan S Alkuraya, Lama AlAbdi, Nour Elkhateeb, Laila Selim, Siddharth Srivastava, Danny D. Nedialkova, Laurence Drouard, Christophe Romier, Efil Bayam, Juliette D. Godin

**Affiliations:** IGBMC, Institut de Génétique et de Biologie Moléculaire et Cellulaire, F-67400 Illkirch, France; CNRS, Centre National de la Recherche Scientifique, UMR 7104, F-67400 Illkirch, France; INSERM, Institut National de la Santé et de la Recherche Médicale, UMR-S 1258, F-67400 Illkirch, France; Université de Strasbourg, IGBMC UMR 7104- UMR-S 1258, F-67400 Illkirch, France; Institut de biologie moléculaire des plantes, CNRS, Université de Strasbourg, 12 rue du Général Zimmer, 67084 Strasbourg, France; Max Planck Institute of Biochemistry, 82152 Martinsried, Germany; Program in Medical and Population Genetics, Broad Institute of MIT and Harvard, Cambridge, MA, USA; Department of Neurology, Boston Children’s Hospital, Boston, MA, USA; Rosamund Stone Zander Translational Neuroscience Center, Boston Children’s Hospital, Boston, MA, USA; Department of Neurosciences, University of California, San Diego, La Jolla, USA; Rady Children’s Hospital, Rady Children’s Institute for Genomic Medicine, San Diego, La Jolla, USA; Department of Pediatrics, School of Medicine, Mashhad University of Medical Sciences, Mashhad, Iran; Department of Medical Genetics, Faculty of Medicine, Mashhad University of Medical Sciences, Mashhad, Iran; Genetic Foundation of Khorasan Razavi, Mashhad, Iran; Division of Genetics and Genomics, Boston Children’s Hospital, Boston, MA, USA; Child Neurology Residency Training Program, Boston Children’s Hospital, Boston, MA, USA; Department of Translational Genomics, Center for Genomic Medicine, King Faisal Specialist Hospital and Research Center, Riyadh 11564, Saudi Arabia; Department of Medical Genomics, Centre for Genomic Medicine, King Faisal Specialist Hospital and Research Center, Riyadh 11564, Saudi Arabia; Neuroscience Center, King Faisal Specialist Hospital and Research Center, Riyadh 11564, Saudi Arabia; Department of Pediatrics, King Faisal Specialist Hospital & Research Centre, Riyadh 11564, Saudi Arabia; College of Medicine, Alfaisal University, Riyadh 11533, Saudi Arabia; Department of Pediatrics, King Faisal Specialist Hospital and Research Center, Jeddah 23433, Saudi Arabia; Department of Pediatrics, Prince Sultan Military Medical Center, Riyadh 12233, Saudi Arabia; Seha Virtual Hospital, Ministry of Health, Riyadh 12382, Saudi Arabia; Department of genetic and metabolic, King Fahad specialist hospital, Dammam 32253, Saudi Arabia; Department of Pediatrics, Pediatric Neurology Unit, King Salman Medical City, Madinah, Saudi Arabia; Department of Pediatrics, Pediatric neurology and metabolic medicine unit, Cairo University, Cairo, Egypt; Department of Pediatrics, College of Medicine, King Fahd Hospital of the University, Imam Abdulrahman Bin Faisal University, Dammam, Saudi Arabia; Department of Pediatrics and Neurology, Harvard Medical School, Boston, MA, USA; Howard Hughes Medical Institute, Boston Children’s Hospital, Boston, MA, USA; Lifera Omics, Riyadh, Saudi Arabia; Department of Clinical Genetics, Cambridge University Hospitals NHS Foundation Trust, Cambridge, UK; Department of Bioscience, TUM School of Natural Sciences, Technical University of Munich, Garching, Germany

**Author notes:** These authors contributed equally to this work (Efil Bayam, Juliette D. Godin).

## Abstract

The ADAT2/ADAT3 complex catalyzes the adenosine to inosine modification at the wobble position of eukaryotic tRNAs. Mutations in *ADAT3*, the catalytically inactive subunit of the ADAT2/ADAT3 complex, have been identified in patients presenting with severe neurodevelopmental disorders (NDDs). Yet, the physiological function of ADAT2/ADAT3 complex during brain development remains totally unknown. Here we showed that maintaining a proper level of ADAT2/ADAT3 catalytic activity is required for correct radial migration of projection neurons in the developing mouse cortex. In addition, we not only reported 20 new NDD patients carrying biallelic variants in *ADAT3* but also deeply characterized the impact of those variants on ADAT2/ADAT3 structure, biochemical properties, enzymatic activity and tRNAs editing and abundance. We demonstrated that all the identified variants alter both the abundance and the activity of the complex leading to a significant decrease of I_34_ with direct consequence on their steady-state. Using *in vivo* complementation assays, we correlated the severity of the migration phenotype with the degree of the loss of function caused by the variants. Altogether, our results indicate a critical role of ADAT2/ADAT3 during cortical development and provide cellular and molecular insights into the pathogenicity of ADAT3-related neurodevelopmental disorder.

## Introduction

Cellular homeostasis and growth require protein synthesis to be both efficient to guarantee sufficient production and accurate to prevent generation of defective or unstable proteins. Efficient and faithful protein translation rely on the activity of transfer RNAs (tRNAs), the adaptor molecules needed to decode genetic information into a peptide sequence. To be fully active, tRNA molecules need to be heavily modified post-transcriptionally. About 30 chemical modifications have been identified at various positions in human tRNAs.^1–3^ On average, a single tRNA carries 13 modifications.^4^ These modifications are catalyzed by different classes of tRNA modifying enzymes and influence tRNA structure, function and stability.^5^ Nucleotides in the anticodon loop are extensively modified.^5^ Those modifications are crucial, as they regulate the tRNA-mRNA interaction to either stabilize cognate Watson-Crick base pairing (position 37) or to facilitate wobble pairing (position 34) to increase the decoding capacity and to prevent frameshift errors.^5^ Thanks to the recent identification of human homologs for many tRNAs modifying enzymes and to the wide use of whole exome sequencing, an increasing number of genes encoding for tRNA modifying enzymes have been linked to human diseases.^3^ Interestingly neurodevelopmental anomalies are the primary manifestation of variants in genes coding for tRNA-modifying enzymes suggesting a specific sensitivity of the brain during development to perturbations in tRNA modifications.^6^ Although most of those variants have been shown to affect tRNAs modification *in vitro*, their direct implication in disease and the underlying pathophysiological mechanisms have only been identified for a few of them.^7–17^

ADAT2/ADAT3 is a heterodimeric enzyme complex that edits adenosine (A) to inosine (I) at the wobble position 34 of tRNAs starting with an A in their anticodon (ANN-tRNAs).^18^ It recognizes specifically tRNAs through the ADAT3 N-terminal domain (ADAT3N), which subsequently rotates to present the bound tRNAs to the ADAT catalytic domain, composed of the C-terminal domain of ADAT3 (ADAT3C) and of ADAT2, that carries the catalytic activity.^19–21^ Given the ability of inosine to pair with uracil (U), cytosine (C) or adenosine (A),^22^ the A to I conversion at position 34 provides an extended base pairing capacity to the modified tRNAs and is essential for decoding the C-ending codons, as GNN-tRNAs do not exist in eukaryotic genomes.^23,24^ In accordance, complete deletion or loss of activity of this tRNA modification complex leads to lethality.^18,25–29^ Knocking down the activity of the complex leads to impaired cell cycle progression^30^ and growth retardation^25^ in several species including yeast and human, possibly due to its role in controlling translational kinetics.^31^

Reflecting a key role of I_34_ modification in brain development, pathogenic variants in *ADAT3* have been identified in patients with neurodevelopmental disorders (NDDs). The same homozygous *ADAT3* variant (NM_138422.4: c.430G>A; p.Val144Met) has been reported in 55 patients from 29 consanguineous families presenting with an autosomal recessive syndromic form of intellectual disability (ID) characterized by developmental delay, moderate to severe ID, speech delay, microcephaly, abnormal brain structure, facial dysmorphism and epilepsy (**Table 1**)^32–38^. In addition to this founder mutation, a homozygous duplication in *ADAT3* (NM_138422.4: c.99_106dupGAGCCCGG; p.Glu36Glyfs*44)^39^ and two compound heterozygous missense *ADAT3* variants in the conserved noncatalytic deaminase domain (NM_138422.4: c.587C>T, p.Ala196Val; c.586_587delinsTT, p.Ala196Leu)^40^ and (NM_138422.4: c.587C>T, p.Ala196Val; c.820C>T p.Gln274*)^17^) have been described in 6 patients with similar but milder ID syndrome features. p.Val144Met variant alters the tRNAs A_34_ deaminase activity of the ADAT2/ADAT3 complex^16,21^ possibly through impaired presentation of ADAT2/3 bound tRNAs to the catalytic site without compromising the formation of the complex.^21^ Yet the structural and functional impact of other variants on ADAT2/3 complex is unknown. Moreover, although I_34_ levels were shown to be decreased in total tRNAs isolated from patients carrying the homozygous p.Val144Met^16^ and compound heterozygous p.Ala196Val; p.Gln274* variants^17^, our knowledge of the molecular effect of ADAT3 variants on specific ADAT-target tRNAs is very limited and the neurodevelopmental processes that require proper function of ADAT2/ADAT3 complex haven’t been elucidated.

**Table 1.**
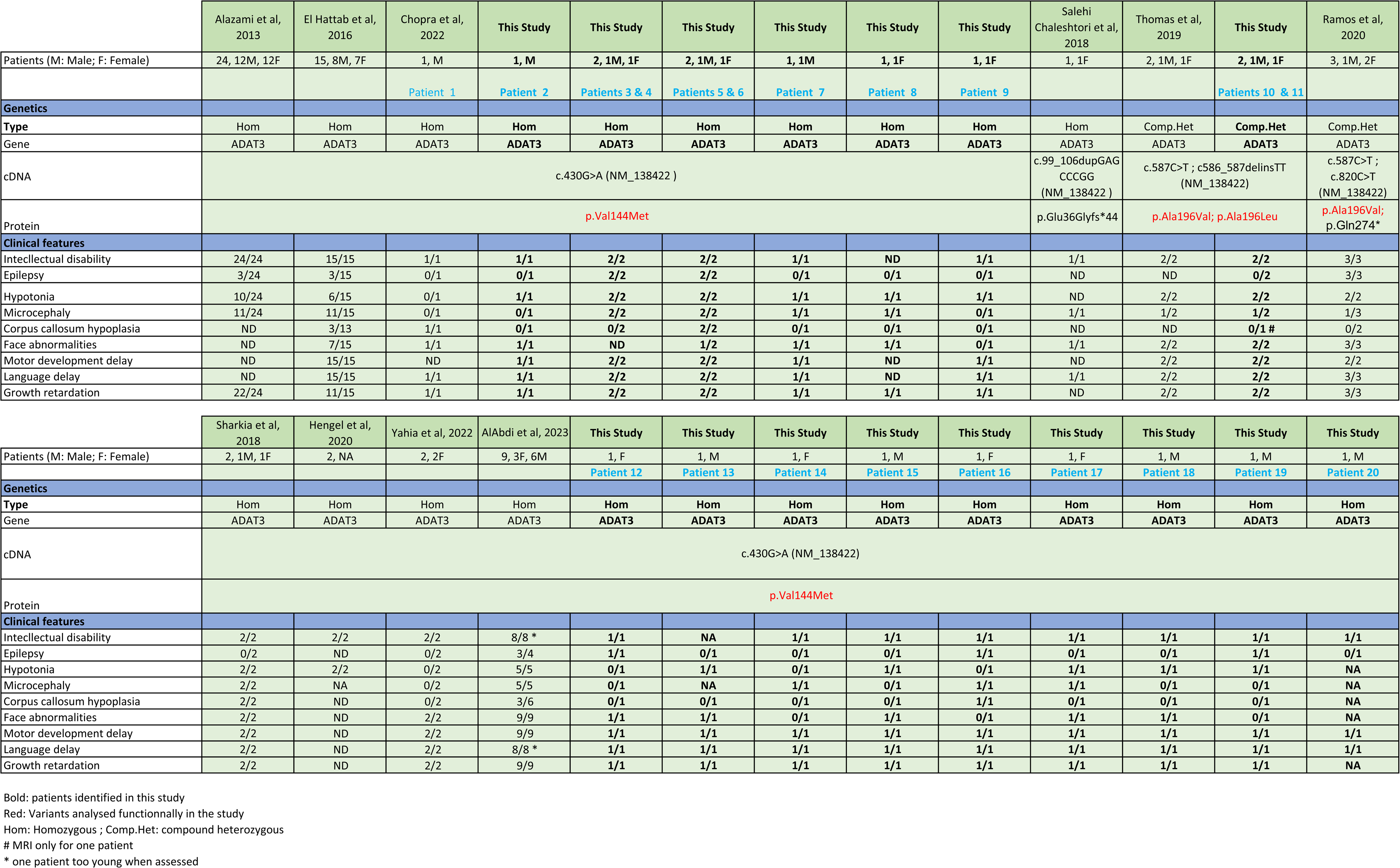
Clinical summary of patients with *ADAT3* variants. Patients identified in the current study are highlighted in bold. *In vivo* functional tests have been performed for the variants depicted in red. ND: not determined; Hom: Homozygous; Comp.Het: Compound heterozygous.

Here we show that the catalytic activity of the heterodimeric ADAT2-ADAT3 complex is critical to promote the radial migration of projection neurons during corticogenesis. We also expand the molecular spectrum of *ADAT3*-related neurodevelopmental disorders by reporting 20 new patients presenting with intellectual disabilities, structural brain anomalies and global growth retardation carrying the previously identified homozygous p.Val144Met variant or the biallelic p.Ala196Val/p.Ala196Leu variants. Using structural, biochemical, molecular and *in vivo* complementation assays, we showed that, although all the variants act through a loss of function mechanism, they have various effects on complex structure, stability and deamination activity that dictate their ability to restore migration defects upon *Adat3* deficiency. We further drew an exhaustive list of tRNA species affected in disease condition, providing strong evidence of a causal relationship between variants in *ADAT3*, loss of translationally competent ANN tRNAs and neurodevelopmental disorders.

## Results

### ADAT2/ADAT3 complex is expressed ubiquitously during cortical development

We first examined the expression pattern of both catalytic and non-catalytic subunits of the ADAT2/ADAT3 heterodimeric complex during mouse cortical development. Although the levels of *Adat3* and *Adat2* mRNA transcripts tend to increase from embryonic day (E) 12.5 to E18.5 **(Fig. 1A)**, immunoblotting using homemade antibodies for ADAT3 and ADAT2 showed rather stable expression of both proteins **(Fig. 1B)**. Immunolabelling of E18.5 embryo brain sections revealed both cytoplasmic and nuclear localization of ADAT3 and ADAT2 in progenitors and neurons **(Fig. 1C-D insets)**. Further analysis of subcellular localization of each protein in day *in vitro* (DIV) 0 and DIV2 primary cortical neurons ascertained the diffused expression pattern of the ADAT2/ADAT3 complex **(Fig. 1E)**.

**Figure 1.**
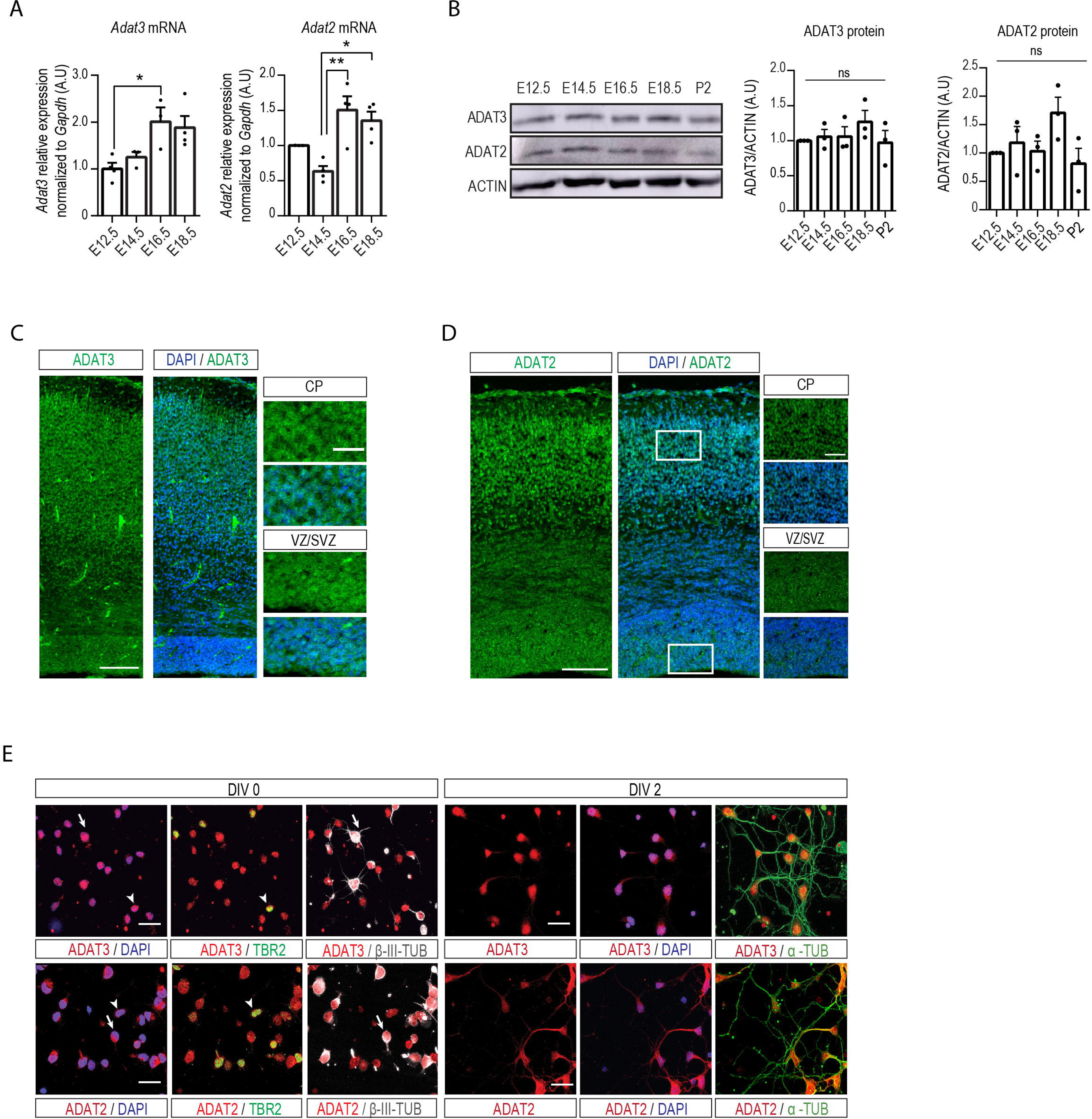
ADAT2/ADAT3 expression pattern in the mouse embryonic cerebral cortex. (**A**) RT-qPCR and (**B**) Western-Blot analysis performed on WT mouse cortices showing expression of *Adat3* and *Adat2* transcripts (**A**) and proteins (**B**) levels throughout development from E12.5 to P2 (n=3-5 cortices per stage). Data are represented as means ± S.E.M and normalized to E12.5. Significance was calculated by one-way ANOVA (Bonferroni’s multiple comparisons test), ns non-significant; *P < 0.05; **P < 0.01. (**C- D**) E18.5 (**C**) and E16.5 (**D**) mouse forebrain coronal sections immunolabelled for (**C**) ADAT3 (green) and (**D**) ADAT2 (green) and counterstained with DAPI (blue) revealing expression of ADAT3 and ADAT2 in both progenitors and neurons. Close-up views of the white boxed area in CP and VZ/SVZ ubiquitous localization of ADAT3 and ADAT2 in the cytoplasm and nucleus. CP cortical plate, SVZ subventricular zone, VZ ventricular zone. Scale bars, 100 μm and 50 μm for magnifications. (**E**) Cortical neurons immunostained for ADAT3 (red), ADAT2 (red), TBR2 (green), α-TUBULIN (α-TUB, green) and ß-III-TUBULIN (ß-III-TUB, gray) and counterstained with DAPI (blue) at 0 or 2 days in vitro (DIV). Arrows point to neurons (cells positive for β-III-TUBULIN). Arrowheads point to intermediate progenitors (cells positive for TBR2). Scale bars, 25 μm.

### ADAT3 regulates radial migration of projection neurons

To evaluate the function of mouse ADAT3 (mADAT3), we cloned two different microRNAs targeting *Adat3* mRNA and confirmed their efficacy by RT-qPCR and immunoblotting (reduction of 85.3% and 96.5% of protein levels for miR1- and miR2-*Adat3* respectively) **(Supplementary Fig. 1A**-B). We first assessed the consequences of acute depletion of m*Adat3* on neuronal migration in wild-type mouse cortices using *in utero* electroporation (IUE) at E14.5 of miRNAs under the control of a ubiquitous CAG promoter together with a NeuroD-IRES-GFP reporter construct, allowing the expression of GFP specifically in postmitotic neurons. Four days after IUE, the distribution of GFP+ neurons depleted for *Adat*3 was significantly impaired with a notable reduction of GFP+ neurons reaching the upper cortical plate (up CP) upon acute depletion of *Adat3* (-32,7% and -21,7 % for miR1- and miR2-*Adat*3, respectively) **(Supplementary Fig. 1C-D).** Yet, after birth, most of the *Adat3*-silenced cells showed a correct positioning with nearly all cells found in the upper layer of the cortex, indicating a delay in migration rather than a permanent arrest **(Supplementary Fig. 1E)**. As pCAGGS is a ubiquitous promoter, impaired neuronal positioning observed upon pCAGGS-driven *Adat3* deletion might result from defects arising in progenitors, in their neuronal progeny or in both. IUE of plasmids expressing the same miRNAs under the control of the neuronal promoter NeuroD showed faulty migration of *Adat3*- silenced neurons with a reduction of 21.9% and 22.7% of cells distributed in the upper CP for miR1- and miR2-*Adat3* respectively, suggesting that defect in postmitotic neurons largely contributed to the *Adat3*- dependent migration phenotype in a cell-autonomous manner **(Fig. 2A-B).** To further validate the specificity of the migratory phenotype induced by *Adat3* silencing, we tested the ability of wild-type (WT) mADAT3 protein to restore the migration defects. We performed co-electroporation of NeuroD-driven miRNAs together with plasmids expressing miRNAs- insensitive mADAT3 under the regulation of a neuronal promoter DCX **(Supplementary Fig. 1F)**. While co-electroporation of WT ADAT3 alone failed to rescue the impaired distribution of *Adat3*-depleted neurons **(Supplementary Fig. 1G**-H), *Adat3*-silenced neurons expressing both wild-type ADAT3 and ADAT2 displayed a correct positioning within the upper cortical plate **(Fig. 2C-D, Supplementary Fig. 1G**-H) indicating that the ability of ADAT3 to restore the migration phenotype depends on the stoichiometric expression of ADAT3 and ADAT2 *in vivo*.^16,18^ Of note, ND-ADAT2, alone **(Supplementary Fig. 1G**-H) or in combination with DCX-ADAT3 **(Supplementary Fig. 5B**-C) did not induce migration phenotypes while overexpressed under control conditions (sh-Scramble). Altogether, these results demonstrate that ADAT3 is cell-autonomously required for proper migration of projection neurons and suggest a contribution of tRNAs modification to radial migration.

**Figure 2.**
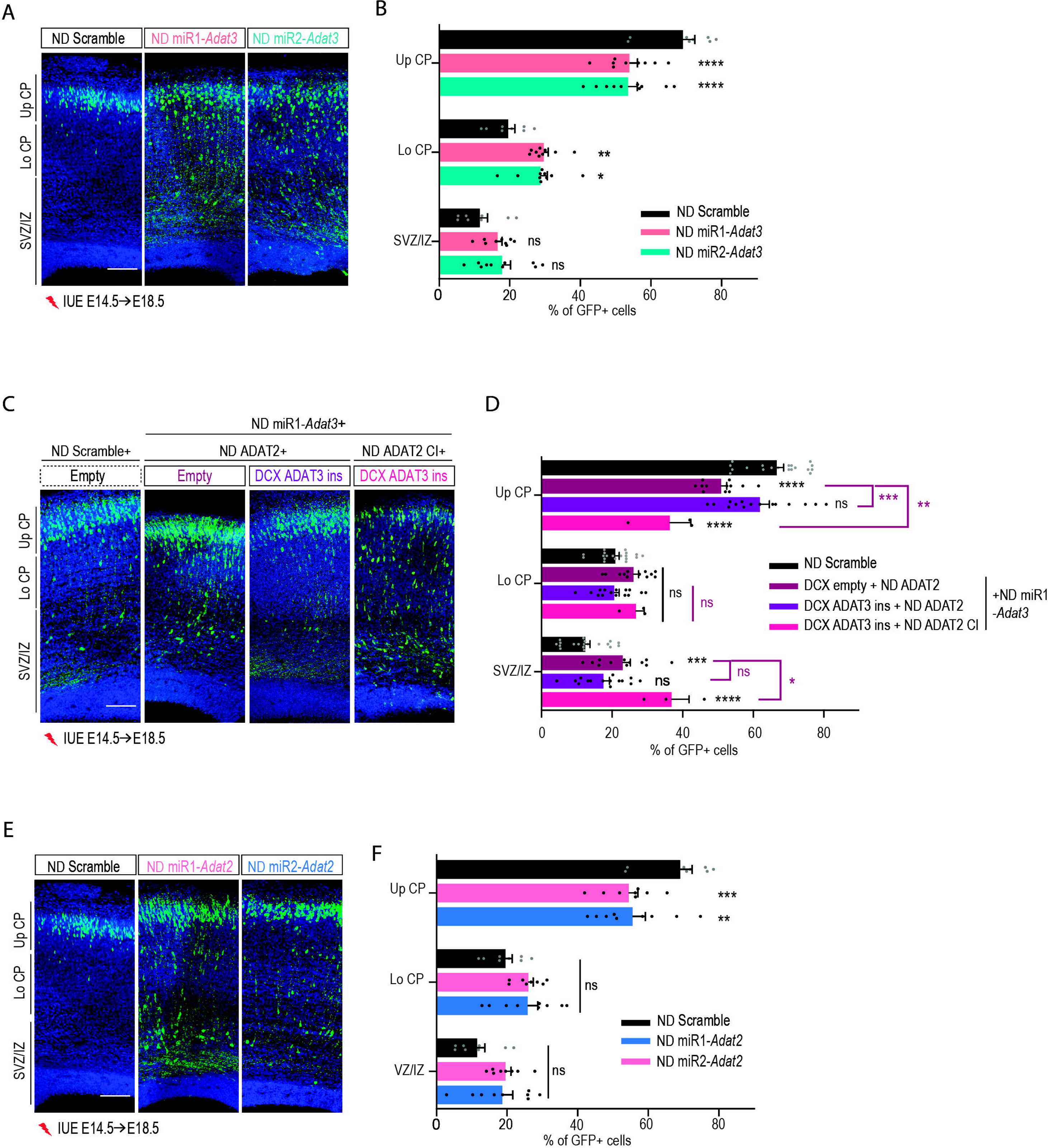
Role of ADAT3 in migrating neurons depends on its function within the ADAT2/ADAT3 complexes. **(A)** Coronal sections of E18.5 mouse cortices electroporated at E14.5 with NeuroD (ND) scramble or two distinct ND-*Adat3* miRNAs (miR1 and miR2) together with ND-GFP. **(B)** Percentage (means ± S.E.M.) of the positive electroporated cells (GFP+, green) in upper (Up CP) and lower (Lo CP) cortical plate, intermediate (IZ) and subventricular zone (SVZ) showing the faulty migration of *Adat3*-silenced neurons. **(C)** Coronal sections of E18.5 mouse cortices electroporated at E14.5 with ND scramble or ND-*Adat3* miR1 together with empty vector or DCX WT ADAT3 (miR1-insensitive (ins)) and WT or catalytically inactive (CI) ND-ADAT2. **(D)** Percentage (means ± S.E.M.) of the positive electroporated cells in upper (Up CP) and lower (Lo CP) cortical plate, intermediate (IZ) and subventricular zone (SVZ) showing the need of catalytic- active ADAT2 for ADAT3 to rescue the faulty migration of *Adat3*-silenced neurons. **(E)** Coronal sections of E18.5 mouse cortices electroporated at E14.5 with ND scramble or two distinct ND-*Adat2* miRNAs (miR1 and miR2), together with ND-GFP. **(F)** Percentage (means ± S.E.M.) of the positive electroporated cells (GFP+, green) in upper (Up CP) and lower (Lo CP) cortical plate, intermediate (IZ) and subventricular zone (SVZ) showing the faulty migration of *Adat2*-silenced neurons. **(A, C, E)** GFP-positive electroporated cells are depicted in green. Nuclei are stained with DAPI. Scale bars, 100 μm. **(B, D, F)** Data were analyzed by two-way ANOVA with Bonferroni’s multiple comparisons test. Number of embryos analyzed: **B,** NeuroD Scramble, n = 8; NeuroD miR1-*Adat3*, n=9; NeuroD miR2*-Adat3*, n=10; **D**, NeuroD Scramble, n = 19; Empty+ NeuroD Adat2 + NeuroD miR1-*Adat3*, n=12; NeuroD Adat2 + DCX Adat3 + NeuroD miR1-*Adat3*, n=15; NeuroD Adat2 C.I *+* DCX Adat3 + NeuroD miR1-*Adat3*, n= 3; **F,** NeuroD Scramble and NeuroD miR1- *Adat2*, n=8; NeuroD miR2*-Adat2*, n=9; ns non-significant; *P < 0.05; **P < 0.01; ***P < 0.001; ****P < 0.0001.

### The catalytic activity of the ADAT2/ADAT3 complex is required for proper neuronal migration

To assess whether the migratory function of ADAT3 depends on its function within the heterodimeric ADAT2/ADAT3 enzymatic complex, we next explored the effect of silencing the catalytically active partner, ADAT2, on neuronal migration. We performed acute depletion of *Adat2* specifically in neurons by IUE of NeuroD-driven *Adat*2-miRNAs in wild type cortices at E14.5. The ability of miRNAs to efficiently target *Adat2* was tested by qPCR and immunoblotting (reduction of 83% of protein levels for both miR1- and miR2- *Adat2*) **(Supplementary Fig. 1A**-B). Consistent with a critical role of the complex in the regulation of radial migration, the migration defects observed after depletion of *Adat2* were comparable to those observed after silencing of *Adat3* (-21%, -19.5% of cells reaching the upper cortical plate in miR1- and miR2-*Adat2* respectively) **(Fig. 2E-F)**. We next addressed whether the ADAT2/ADAT3 complex controls neuronal migration through its catalytic activity and tested for rescue of the phenotype induced by the loss of *Adat3* by expressing wild-type ADAT3 and a catalytically inactive form of ADAT2 (ADAT2 CI, **Supplementary Fig. 1I)**.^18,21,41^ In accordance with the need of the enzymatic activity, co-expression of ADAT3 with wild-type ADAT2 but not with catalytic inactive ADAT2 rescued the impaired positioning of *Adat3*-depleted cells **(Fig. 2C-D).** Altogether these results demonstrate the catalytic activity of the ADAT2/ADAT3 complex is required to exert its function in migrating projection neurons.

### Identification of novel patients with *ADAT3* variants

Through the GeneMatcher^42^ and Matchmaker Exchange^43,44^ platforms, we identified 20 individuals from 17 unrelated families carrying biallelic variants in *ADAT3*, presenting with intellectual disabilities (IDs) and brain malformation (**Table 1**). These individuals included 11 males and 9 females with ages ranging from 9- months to 16-years. Of note, Patient 1 has been previously reported in a cohort of 50 probands with cerebral palsy.^33^ Patients 1 to 9 and 12 to 20 carry the homozygous p.Val144Met (p.V144M) variant (NM_138422, c.430G>A), the most common cause of autosomal recessive ID in Arabia, whereas Patients 10 and 11 had the previously identified^40^ compound heterozygous variant p.Ala196Leu(p.A196L)/p.Ala196Val (p.A196V) (NM_138422, c.587C>T; c586_587delinsTT). In accordance with previous reports^17,32–37,39,40^, commonly observed clinical features included global developmental delay (19/19), ID (18/18), muscle tone defects (15/19), microcephaly (10/18), and epilepsy (7/20) (**Table 1**, **Supplementary Table 1**, **Supplementary note 1**). All patients presented with motor delay and language deficit ranging from severely impaired speech to non-verbal. Magnetic resonance imaging (MRI) images were available for 15 patients (Patients 1, 2, 5, 6, 7, 9, 10, 12 to 19). While Patients 7, 10, 12, 13, 14, 15, 17 and 18 showed a normal brain structure, other patients displayed variable brain structural anomalies including dysplastic appearance of the corpus callosum (Patients 1, 5, 6, 16 and 19), microcephaly (Patients 2, 5 and 16), abnormal gyrification (Patients 5 and 6), enlarged ventricles (Patients 5, 6, 16 and 19), cavum septum pellucidum (Patient 9) and nearly absent myelination (Patient 5) (**Fig. 3A-F**). Overall, we expanded the clinical spectrum of *ADAT3* related neurodevelopmental disorders by presenting 20 patients displaying severe neurodevelopmental delay associated with heterogenous brain anomalies (**Fig. 3G**, **Table 1, Supplementary Table 1, Supplementary note 1**). To further interrogate the molecular effect of the *ADAT3* variants, we analyzed the level of expression of ADAT3 in patient’s samples when available. We compared hADAT3 protein levels in lymphoblastoides cell lines (LCLs) homozygous for the p.V144M variant (affected patient - Fam1 in^32^) and p.A196V/p.A196L (Patients 10 and 11) depicted in **Fig. 3E** to control lymphoblasts generated from sex and aged-matched healthy individuals. Though the levels of *ADAT3* transcripts remained stable (**Supplementary** Fig. 2), immunoblotting with two different antibodies revealed that both ADAT3 p.V144M and p.A196V/p.A196L variant lead to a severe but not a total decrease of ADAT3 protein levels (**Fig. 3H**), in line with the total loss of ADAT3 being incompatible with life.^18,25–29^

**Figure 3.**
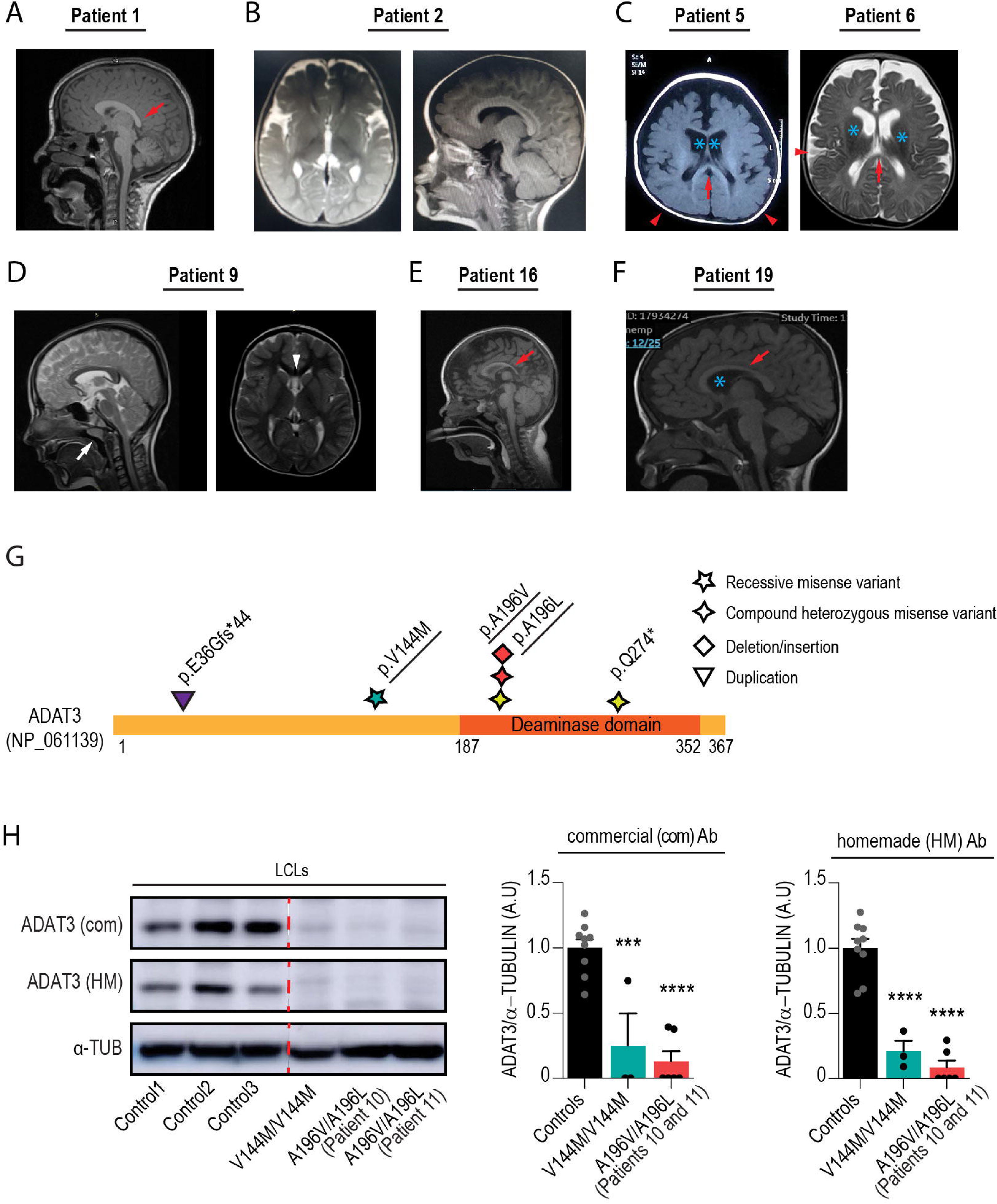
Clinical features of patients with *ADAT3* variants. **(A-F)** Axial and/or sagittal T1 and T2- weighted brain MRI images of (**A**) Patient 1, (**B**) Patient 2, (**C**) Patients 5 and 6, (**D**) Patient 9 (**E**) Patient 16 and (**F**) Patient 19. The red arrows point to thin corpus callosum, red arrowhead indicates the simplified gyral pattern of the cortex. Blue asterisks show enlarged ventricles. White arrow and arrowhead indicate, respectively, adenoid hypertrophy and cavum septum pellucidum. **(G)** Schematic representation of human ADAT3 protein indicating positions of all variants identified so far. Variants depicted in the same color were found in the same patient. *In vivo* functional tests have been performed for the variants that are underlined. **(H)** Western Blot analysis of p.V144M/p.V144M and p.A196V/p.A196L (Patients 10 and 11) patient LCLs revealing reduced ADAT3 protein levels in comparison to controls (Controls, n=9; V144M/V144M, n=3 and A196V/A196L, n=6 (3 of each Patient)). α-TUBULIN (α-TUB) is used as a protein loading control. Both commercial (com) and homemade (HM) antibodies have been used to detect ADAT3 proteins. Red dashed line indicates where the membrane was cut. One-way ANOVA, Bonferroni’s multiple comparisons test. ***P < 0.001; ****P < 0.0001.

### Variants in *ADAT3* affect the stability, structure and enzymatic activity of the ADAT2/ADAT3 complex

We next evaluate the impact of the identified variants on the remaining ADAT2/ADAT3 complex. We first mapped the variants using the crystal structure of the mouse WT ADAT2/ADAT3 complex.^21^ The ADAT3 V128 residue (corresponding to the V144 residue in human, **Supplementary Fig. 3A**) is part of a large hydrophobic core located in the middle of the N-terminal domain of ADAT3 (ADAT3N) ^21^, while the ADAT3 A180 residue (corresponding to the A196 residue in human), is buried within the ADAT3 C-terminal domain (ADAT3C, **Fig. 4A**). Despite their different three-dimensional locations, the p.V144M and p.A196V/L variants cause similar clinical phenotypes raising the question of the impact of these mutations on ADAT3 and ADAT2/ADAT3 complex solubility, structure and activity.

**Figure 4.**
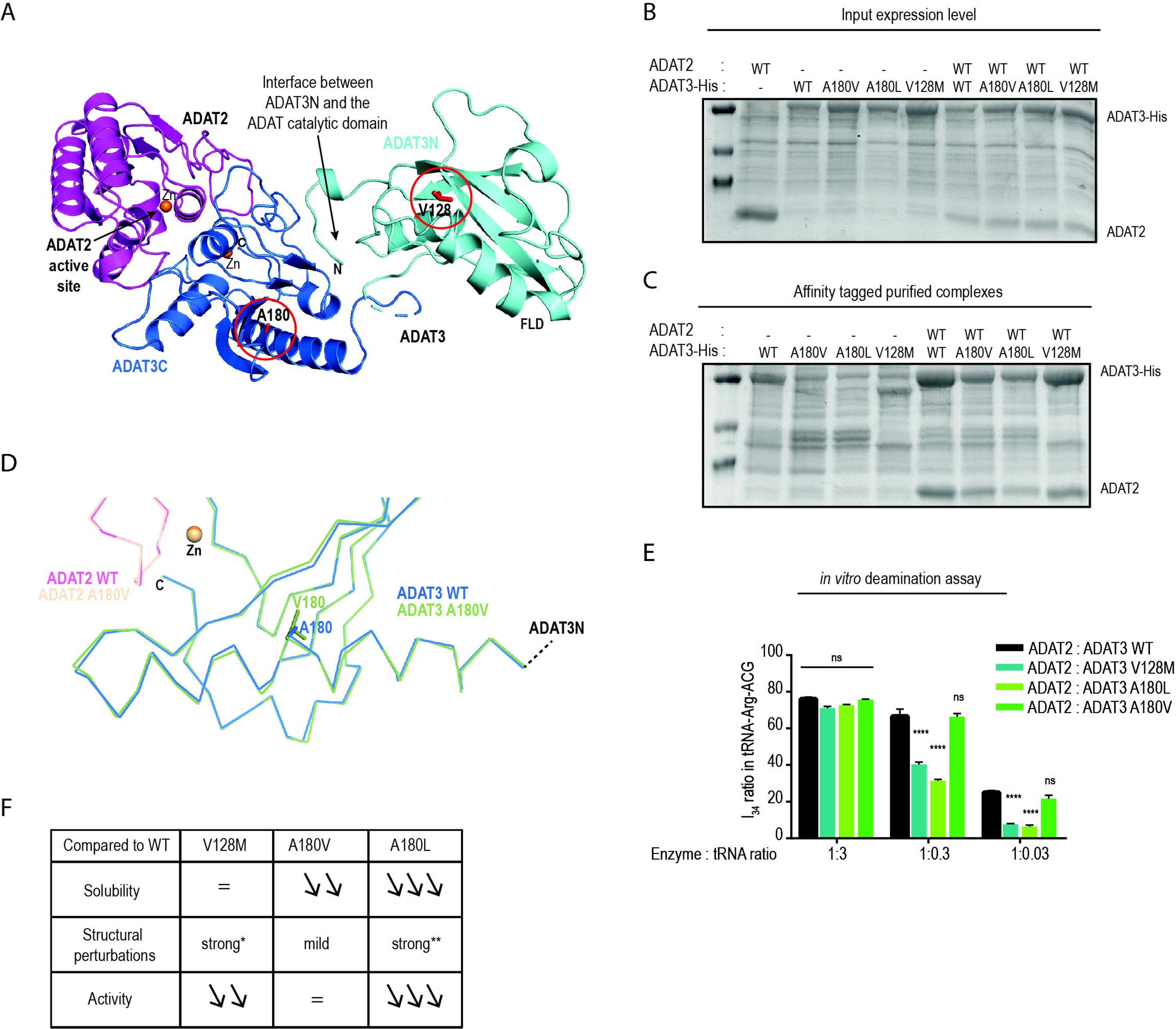
The V128M and A180V/L ADAT3 mutants affect ADAT2/ADAT3 stability, structure and deamination activity. **(A)** Ribbon representation of the crystallographic structure of the WT mouse ADAT complex (PDB entry 7nz8). The catalytic domain of ADAT is composed of ADAT2 (magenta) and of the C-terminal domain of ADAT3 (blue; ADAT3C). The N-terminal domain of ADAT3 (cyan; ADAT3N) is key to recognize tRNAs through its Ferredoxin-like domain (FLD) and to rotate with respect to the ADAT catalytic domain to position the tRNA anticodon loop within the ADAT2 active site. The two residues (V128 and A180), shown in red and that are found mutated in patients, are displayed. These are located in different regions of the ADAT complex. **(B)** SDS-PAGE analysis of expression levels in *E. coli* of untagged ADAT2 WT and His-tagged ADAT3 (WT and A180V, A180L, V128M mutants) constructs, either alone or in combination. Expression levels are similar for all constructs used. **(C)** SDS-PAGE analysis of His-tag affinity-purified samples of (**B**). In the absence of ADAT2, the ADAT3 mutant constructs show a significant decrease in solubility compared to WT. Co-expression of ADAT2 with these constructs restore solubility upon formation of the ADAT2/ADAT3 complex albeit to different extents. **(D)** Close-up view and comparative structural analysis of the region of the ADAT complex harboring A180 in the ADAT2/ADAT3 and ADAT2/ADAT3-A180V complex structures. The A180V mutation induces local changes in the main chain neighboring secondary structure elements, including in the central β-sheet organizing the ADAT3 C-terminal domain. **(E)** Deamination assays for mouse ADAT3 WT, A180V, A180L and V128M in complex with ADAT2. Whereas the WT and A180V complexes have similar activities, the A180L and V128M complexes show a similar decrease in activity. Data (means ± S.E.M) from 3 different experiments per condition were analyzed by two-way ANOVA, with Dunnett’s multiple comparison test. ns non-significant; ****P < 0.0001. (**F**) Summary table showing that the loss of enzymatic activity correlates with structural alterations rather than with impaired solubility. Asterisk and double asterisk indicate prediction based on the structure of the ADAT2/ADAT3-V128L and ADAT2/ADAT3-A180V, respectively.

Bacterial expression of the WT and mutant mouse ADAT3 constructs alone showed that, despite similar expression levels (**Fig. 4B**), all three mutants affect significantly the solubility of ADAT3 (**Fig. 4C**). However, co-expression of these constructs with ADAT2 partially restored ADAT3 solubility upon formation of the ADAT2/ADAT3 complex. Co-expressed with ADAT2, the V128M ADAT3 construct was almost as soluble as the WT construct, whereas the A180V and A180L constructs were less soluble, albeit the A180V being slightly more soluble than the A180L construct (**Fig. 4C**). Interestingly, as assessed during the purification process, once formed and soluble, the mutant complexes do not show aggregation properties and can be used for further biochemical assays and structural analyses. We therefore sought to characterize the structural effect of the A180V/L mutations as it was done previously for the variant at the V128 residue.^21^ In our initial study, crystals have been obtained for the ADAT2/ADAT3-p.V128M complex but those did not diffract sufficiently. However, the structure of the ADAT2/ADAT3-V128L complex showed that the V128L mutation perturbs the ADAT3N region involved in the interaction with the ADAT catalytic domain, suggesting a partially impaired presentation of the bound tRNA anticodon loop to the ADAT2 active site. These defects should be exacerbated in the case of the V128M mutant.^21^ Crystallization assay of the ADAT2/ADAT3-A180V and ADAT2/ADAT3-A180L complexes were successful for the former complex, while the latter failed to crystallize. Following structure determination at 2.9 Å resolution (**Supplementary Table 2**), comparison with the structure of the WT ADAT2/ADAT3 complex revealed that, unlike the p.V128L variant that affects mostly the ADAT3N, the p.A180V substitution causes small local perturbations in the structure of the ADAT3 C-terminal domain (**Fig. 4D**). We anticipate in this case also that this effect is exacerbated with the A180L mutant due to the bulkier character of the leucine residue compared to the valine residue. The structural effects caused by the mutation of the A180 residue are more difficult to predict than those on the V128 residue.^21^ Indeed, this residue is relatively far away from ADAT2 and its active site, and it is unlikely that the mutation of this residue directly affects the catalytic mechanism. On the other hand, A180 is located in the long α-helix that immediately follows the ADAT3N domain (**Fig. 4A**). Its mutation into leucine could affect the proper folding and positioning of this helix but also those of the central ADAT3 C-terminal domain β-sheet that is also involved in the interaction with ADAT3N. Therefore, our structural analysis suggested that the A180L mutation probably also affects the interaction between the ADAT3N domain and the ADAT catalytic domain, thereby hampering the correct presentation of the tRNA to the ADAT catalytic domain and, consequently, the deaminase activity.

To ascertain the effect of the variants on the enzymatic activity of the ADAT2/ADAT3 complex, we performed sequencing of an *in vitro*-transcribed cognate tRNA, tRNA Arg(ACG), after incubation with different amounts of purified recombinant WT or mutant ADAT2/ADAT3 complexes.^21^ As inosine is read as a ‘G’ by reverse transcriptase,^28^ we sought for the percentage of G_34_ as a proxy of A_34_ to I_34_ editing. While the p.V128M ADAT3 exhibited a significantly impaired enzymatic activity (-68 % of A_34_ to I_34_ with the lowest concentration of the ADAT2/ADAT3-V128M- complex) as expected,^16,21^ the ADAT2/ADAT3-A180V heterodimer retained a comparable activity to the WT complex and the ADAT2/ADAT3-A180L displayed a severely diminished deamination capacity (-76% compared to the WT complex at the lowest concentration) (**Fig. 4E, Supplementary Fig. 3B**). Notably, the two variants found in the Patients 10 and 11 (p.A180V/L; corresponding to p.A196V/L in human) impair ADAT2/ADAT3 deamination activity differently, although they affect the solubility of the ADAT2/ADAT3 complex similarly (**Fig. 4C**). Along the same line, the p.V128M ADAT3 variant strongly affected the enzymatic activity of the complex despite any effect on its solubility (**Fig. 4C,E**). Overall, these data strongly suggest that the level of deamination activity does not directly correlate to the level of solubility but rather indicate that the faulty deamination activity of the mutant complexes might mostly stem from structural perturbations (**Fig. 4F**).

### Selective loss of I_34_ modification and reduced steady state of cognate tRNAs in patient derived cells

Given that ADAT3 variants have various effects on the structure, stability and enzymatic activity of the ADAT2/ADAT3 complex, we next investigated whether the impaired ADAT2/ADAT3 function affects the A_34_ to I_34_ tRNA editing. We took advantage of the recently developed mim-tRNA-seq method, that allows robust quantification of individual tRNA species as well as determination of presence and stoichiometry of 8 tRNA modifications, including inosine.^45,46^ In our dataset, >72% of the reads were unique and mapped at 95% on average to nuclear-encoded tRNAs (**Supplementary Fig. 4A**-B). More than 67% of the uniquely mapped tRNAs were full length tRNAs of which >97% contained the 3’CCA tail indicating that they were mature, translationally competent tRNAs (**Supplementary Fig. 4C**-D). We first compared the I_34_ proportion in patient and control LCLs at the level of isodecoders, defined as tRNA transcripts sharing the same anticodon but differing elsewhere in their sequence. The isodecoders of the eight ANN tRNA families (Ala-AGC, Arg-ACG, Ile-AAU, Leu-AAG, Pro-AGG, Ser-AGA, Thr-AGU and Val-AAC) are fully deaminated in control condition, as previously described in other cellular contexts^18,47,48^ (**Fig. 5A-B, Supplementary Table 3**). However, we observed a consistent deamination defect in both p.V144M/p.V144M and p.A196V/p.A196L patient cells. While isodecoders belonging to the tRNA-Arg-ACG, tRNA-Pro-AGG and tRNA-Ser-AGA families were not affected, the other tRNAs showed a substantial decrease in I_34_ proportion, the tRNA-Ala-AGC family being the most affected with 4 isodecoders out of the 6 detected showing a decreased A_34_ to I_34_ editing (mean of 43%, 13%, 31% and 79% of I_34_ for, respectively, Ala-AGC-1 Ala-AGC- 3, Ala-AGC-4, and Ala-AGC-11 in the 3 mutant cell lines) (**Fig. 5A-B, Supplementary Table 3**). As a result, when grouped by anticodon pools (see material and methods), we observed a 16-25% decrease of deamination for Ala-AGC in both mutant conditions compared to the control, while we did not detect any changes in the level of I_34_ for the other 7 ANN anticodon families (**Supplementary Fig. 4E**-F**, Supplementary Table 3**). Of note among the other modifications that could be identified by mim-tRNA-seq (m^1^A, m^1^G, m^2^_2_G, m^3^C, yW, acp^3^U, I, m^1^I, m^3^C_32_ and m^1^A_58_) in some isodecoders showed very slight decrease in the patient derived cell lines compared to controls, confirming the specificity of the I_34_ perturbation and the lack of interdependence of I_34_ with these other detected modifications (**Fig. 5C**).

**Figure 5.**
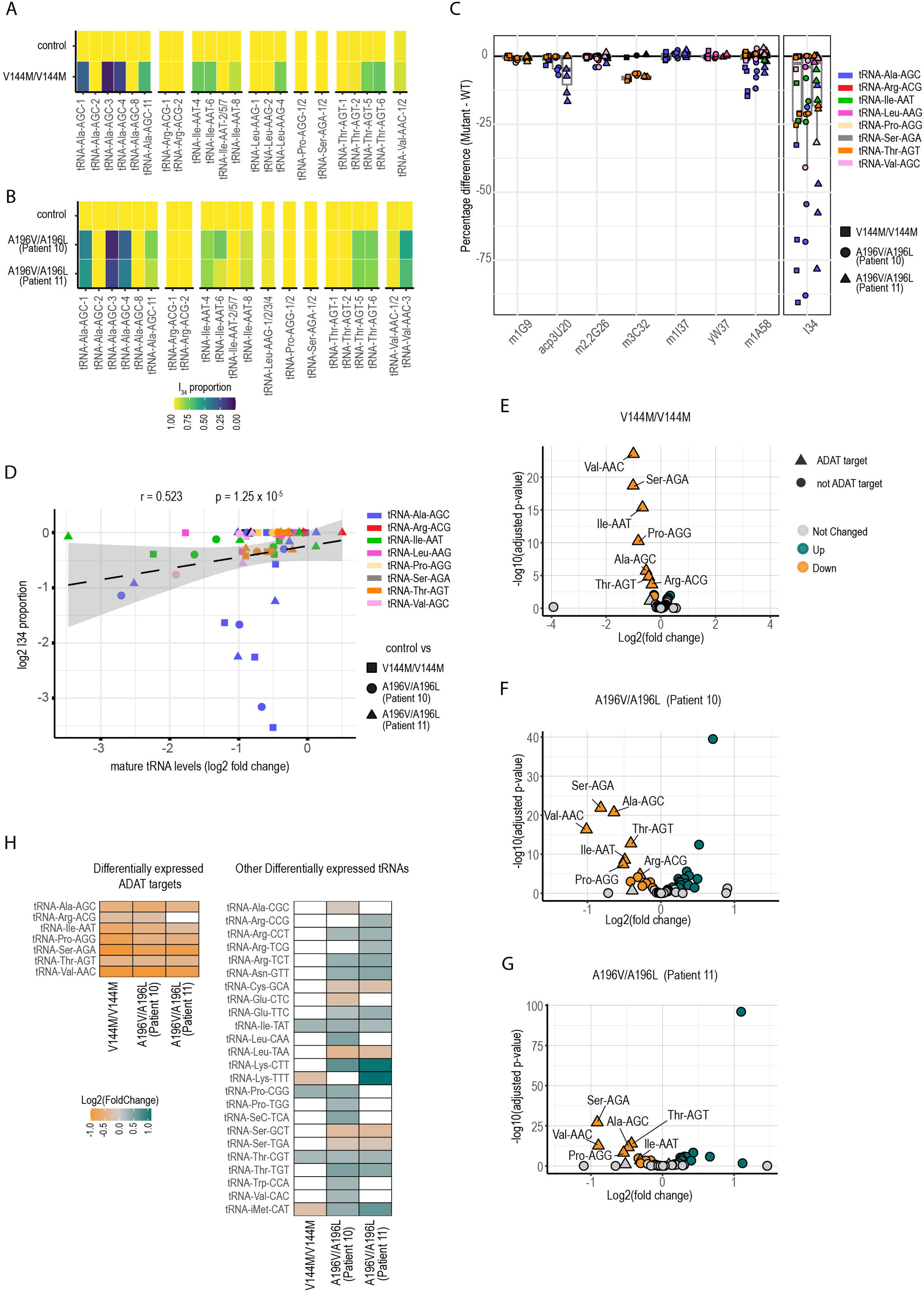
Deamination and abundance of ADAT2/ADAT3 target tRNAs are decreased in patient cells. (A-B) Heatmap showing I_34_ levels in ADAT target tRNA isodecoders in LCLs derived from (**A**) p.V144M/ p.V144M and (**B**) p.A196V/p.A196L patients compared to control. (Controls, n=2; V144M/V144M, n=2; A196V/A196L (Patient 10) n=3 ; A196V/A196L (Patient 11) n=3). (**C**) Graph showing the percentage change in ADAT target tRNAs of all other 7 modifications (m^1^G9, acp^3^U20, m^2^_2_G26, m^3^C32, m^1^I37, yW37, m^1^A58) that can be detected by mim-tRNA seq. Change at I_34_ levels is depicted as a side view for comparison. tRNA isoacceptors from comparison of control LCL to LCLs derived from V144M/V144M, A196V/A196L (Patient 10) and A196V/A196L (Patient 11) are depicted with square, circle and triangle respectively. Isoacceptors are coloured as indicated. (**D**) Graph showing the correlation between the change in I_34_ proportion and the change in mature tRNA levels in log2 scales. Black dashed line is the trend line and standard deviation is shown with a grey zone. tRNA isodecoders from comparison of control LCLs to LCLs derived from V144M/V144M, A196V/A196L (Patient 10) and A196V/A196L (Patient 11) are depicted with square, circle and triangle respectively. Isodecoders are coloured as indicated. **(E-G)** Volcano plot showing the negative log_10_ adjusted *P*value (p-adj) of all tRNAs pooled at the anticodon level against their log_2_ fold change (log_2_FC) in LCLs derived from (**E**) p.V144M/p.V144M (n=2), (**F**) p.A196V/p.A196L (Patient 10, n=3) and (**G**) p.A196V/p.A196L (Patient 11, n=3) compared to control (Controls, n=2). Triangle and circle show ADAT targets and non-target tRNAs respectively. Green, orange and grey represent upregulated, downregulated and unchanged tRNAs respectively based on DESeq2 padj<0.05. (**H**) Heatmap showing log2 DESeq2 fold change of differentially regulated ADAT target tRNAs and non-target tRNAs summed by anticodon. White boxes show non-significant ones.

We next used mim-tRNA-seq data to compare the abundance of mature tRNAs in control and ADAT3 mutant LCLs. Among the 379 cytoplasmic tRNAs isodecoders for which we reached a single-transcript resolution (90% of the predicted cytoplasmic tRNAs), 19%, 25%, and 28% were differentially expressed in p.V144M/p.V144M and in the two p.A196V/p.A196L (Patients 10 and 11) mutant cells compared to control respectively (adjusted P (Padj) ≤ 0.05) (**Supplementary Fig. 4G**-I**, Supplementary Table 3).** Interestingly, up to 37.5% of the differentially expressed tRNAs are ADAT2/ADAT3 substrates with 57% of all ANN isodecoders being deregulated versus 30% of non-INN tRNAs transcripts (**Supplementary Table 3**). These data were highly reproducible among replicates as shown in the principal component analysis (**Supplementary Fig. 4J**). Strikingly, we observed a significant inverse correlation (Spearman’s correlation coefficient r=0.523, p=1.25x10^-5^) between the decrease in cellular abundance of mature tRNA isodecoders and their level of deamination (I_34_) (**Fig. 5D**), indicating that the stability of ANN isodecoders is very sensitive to the loss of I_34_. We then aggregated all tRNAs by their anticodon and reperformed a differential expression analysis. We showed that, although some anticodon families that are not targets of the ADAT2/ADAT3 complex were deregulated (**Fig. 5E-G**, circle points, **Supplementary Fig. 4K**), 6 out of the 8 tRNAs anticodon families, that showed a decreased expression of up to 2-fold in all three mutant cells lines relative to control cells, were ANN tRNAs (**Fig. 5H**). Of note, the two other non-ANN tRNA commonly deregulated were up-regulated (Thr-CGT and lle-TAT).

Overall, these results indicate that the combined decreased expression of the ADAT2/ADAT3 complex and diminished activity of the remaining complexes observed in ADAT3 mutant conditions significantly change the cellular pools of ANN tRNAs, that likely stem from both an excessive proportion of non-translationally competent A_34_ tRNAs and selective degradation of instable A_34_ tRNAs isodecoders.

### Missense variants in *Adat3* impair neuronal migration through loss of function mechanism

To further assess the functional consequences of *ADAT3* variants and to ascertain the predicted loss of function mechanism, we evaluated the ability of the variants to restore the migration phenotype induced by the depletion of *Adat3*. As we observed a decrease in protein levels in both p.V144M/p.V144M and p.A196V/p.A196L ADAT3 LCLs **(Fig. 3M)**, we first tested whether the rescue of the migration phenotype observed upon m*Adat3* deletion depends on ADAT3 dosage by IUE of miR1-*Adat3* together with increasing amount of mADAT3. Whereas *Adat3*-silenced neurons expressing 1 unit (0.75 μg/μl) of mADAT3 are correctly distributed in the upper cortical plate 4 days after IUE (**Fig. 2C, D**), expression of two-thirds of unit (0.5 μg/μl) of mADAT3 only partially rescued the faulty migration (**Fig. 6A, B**), suggesting that ADAT3 controls neuronal migration in a dose-dependent manner. Next, to undoubtedly demonstrate the loss of catalytic activity of the remaining complexes, we performed complementation assay by expressing p.V128M, p.A180V and p.A180L mADAT3 variants (corresponding to human p.V144M, p.A196V, p.A196L variants, respectively) together with wild-type mADAT2 in *Adat3*-silenced neurons. When co-expressed with ADAT2, all variants showed expression similar to the expression of the wild-type protein and their expression in control condition (Scramble miRNA) did not induce any migration phenotype (**Supplementary** Fig. 5). Interestingly, the p.A180V variant that does not affect the deamination activity of the ADAT complex as demonstrated *in vitro* (**Fig. 4E**) restored the migration defects as efficiently as the ADAT3 WT construct (**Fig. 6A, C**). On the contrary, the p.A180L and p.V128M variants that strongly impair the activity of the ADAT2/ADAT3 complex (**Fig. 4E**), fail to rescue the migration phenotype observed upon *Adat3* depletion (**Fig. 6A, C**). Altogether, these results demonstrate that missense hADAT3 variants impede the radial migration of projection neurons through either loss of expression and/or catalytic activity.

**Figure 6.**
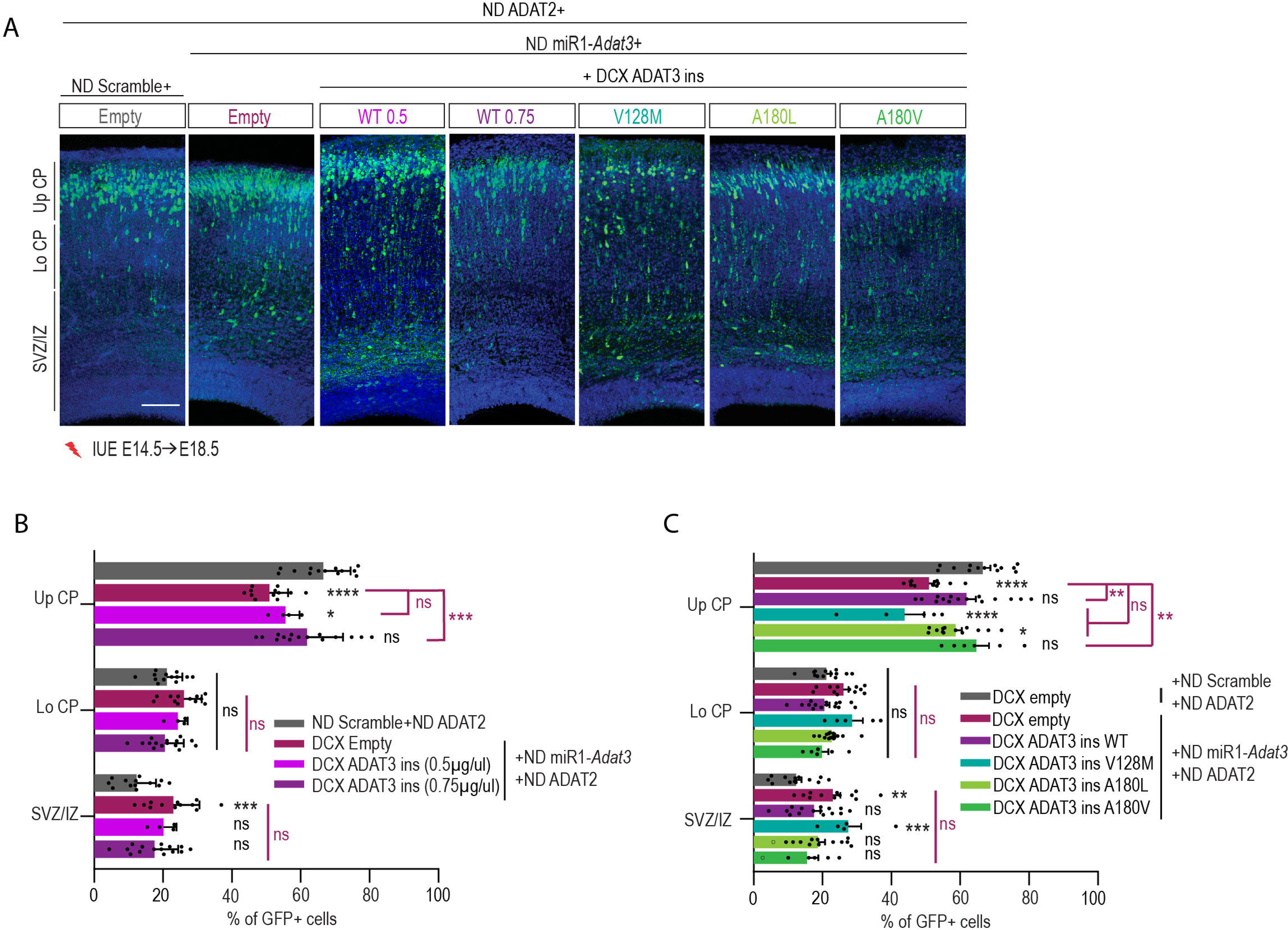
**Missense variants in *Adat3* impair neuronal migration**. **(A)** Coronal sections of E18.5 mouse cortices electroporated at E14.5 with ND ADAT2 and ND-GFP together with either NeuroD scramble or ND *Adat3* miRNAs in combination with DCX Empty; DCX ADAT3 (miR1-insensitive (ins)) at two different concentration (0.5 or 0.75μg/μl) or various variants (at 0.75μg/μl). GFP-positive electroporated cells are depicted in green. Nuclei are stained with DAPI. Scale bar, 100 μm**. (B, C)** Analysis of percentage (means ± S.E.M.) of electroporated cells in upper (Up CP) and lower (Lo CP) cortical plate, intermediate (IZ) and subventricular zone (SVZ) showing a dose-dependent rescue of migration with wild-type proteins and absence of rescue with most of the variants. Data were analyzed by two-way ANOVA (Tukey’s multiple comparison test). Number of embryos analyzed: NeuroD Scramble, n = 13; NeuroD miR1-*Adat3 +* Empty, n=12; NeuroD miR1-*Adat3 +* DCX WT (0.5 μg/μL), n=4; NeuroD miR1- *Adat3 +* DCX WT (0.75 μg/μL), n= 15; NeuroD miR1-*Adat3 +* DCX V128M, n=5; NeuroD miR1-*Adat3 +* DCX A180L, n=13; NeuroD miR1-*Adat3 +* DCX A180V, n=6; ns non-significant; *P < 0.05; **P < 0.01; ***P < 0.001; ****P < 0.0001.

## Discussion

Our findings highlight a critical role of the heterodimeric enzyme complexes, ADAT2/ADAT3, in the regulation of radial migration of projection neurons. We provide several lines of evidence which suggest that the catalytic activity of the ADAT2/ADAT3 complex is required for proper neuronal migration. First, we demonstrate that silencing of both the catalytic (ADAT2) and non-catalytic (ADAT3) subunits of the complex impaired neuronal migration to similar extent (**Fig. 2**). Second, co-expression of ADAT3 together with ADAT2 is required to abolish the phenotype induced by the loss of *Adat3* (**Supplementary Fig. 1G**,H), suggesting that co-abundance of ADAT2 is likely necessary to stabilize ADAT3 *in vivo*. This result correlates with *in vitro* findings showing that ADAT3 tends to self-associate and aggregate when not assembled with ADAT2.^16^ In addition, co-expression of the catalytic-inactive form of ADAT2 and ADAT3 is unable to rescue the *Adat3*-induced migratory phenotype (**Fig. 2E-F**). Third, while the ADAT2/ADAT3 complex bearing the p.A196V (A180V mADAT3) variant, that retained a similar enzymatic activity than the WT ADAT complex, restored the faulty migration observed upon *Adat3* depletion; the ADAT2/ADAT3 complexes bearing, the p.V144M (V128M mADAT3) and the p.A196L (A180L mADAT3) variants respectively which exhibit reduced enzymatic activity lost their ability to rescue the migration phenotype (**Fig. 4E**, **Fig. 6**). Fourth, combined decreased expression and reduced catalytic activity of the ADAT2/ADAT3 complex in patient derived cell lines impaired the A to I conversion at the wobble position of tRNAs, to an extent that is compatible with life but that likely causes detrimental brain phenotypes (**Fig. 5**, **Fig. 6**).

We examined the pathological mechanisms associated to ADAT2/ADAT3 complex related NDD at the genetic, structural/biochemical and molecular levels. All results converged towards a loss of function mechanism. At the genetic level, we expanded the clinical spectrum of ADAT related NDDs by reporting 18 new patients from 16 different families carrying the previously identified^32–37^ p.V144M/pV144M variant and 2 new patients from the same family carrying the previously reported^40^ biallelic p.Ala196Val/p.Ala196Leu variant. Clinical presentation and MRI images of the newly-identified individuals matched with the clinical features observed in patients with ADAT-related NDDs.^17,32–37,39,40^ Interestingly, variants in *ADAT3* gene identified in patients with neurodevelopmental disorders were only found at the biallelic state (**Fig. 3**, **Table 1**). Consistently, human *ADAT3* and *ADAT2* genes tolerate loss-of-function variants with many loss-of-function heterozygous variants reported in the gnomAD general population (Genome Aggregation Database, v4.0.0), suggesting that cortical development can sustain *ADAT3* hemizygosity. Of note, no biallelic null variant have been found in *ADAT2* yet. Corroborating the human findings, we showed that full knock-out of *Adat3* is lethal in mice (data not shown), and that expression of wild-type mADAT3 restored the neurodevelopmental phenotype in a dose-dependent manner (**Fig. 6A-B**). Altogether these findings indicate that a minimal level of complex activity is required to ensure proper mammalian neuronal development.

We further provided insights into the structural basis of the ADAT3-related NDD. Although the V128 (V144 in Human) and A180 (A196 in Human) residues lie in different structural domains, the N-terminal (ADAT3N)^21^ and C-terminal domains respectively, the V128M and A180L variants are both suggested to affect the presentation of the tRNAs anticodon loop to the active catalytic site of the complex, yet through distinct structural perturbation. Based on the crystal structure of the ADAT2/V128L-ADAT3 mouse complex, we previously predicted that the V128M variant hinders the proper positioning of the ADAT3N terminal domain to present correctly the anticodon loop to ADAT2, likely through perturbation of the interaction between ADAT3N and the ADAT2/ADAT3 catalytic domain.^21^ Here, the crystal structure of the ADAT2/A180V-ADAT3 mutant complex did not reveal any defect in the ADAT3N domain, but rather local perturbations in the α-helix, where the A180 residue is located, and in the ADAT3 C-terminal domain β- sheet, both located close to interaction surface with ADAT3N. We anticipated that the A180L substitution aggravates these local perturbations and alters the interaction between the ADAT3N and the ADAT2/ADAT3 catalytic domains and therefore the correct presentation of the tRNAs to ADAT2. This structurally predicted loss of enzymatic activity of the ADAT2/V128M-ADAT3 and ADAT2/A180V-ADAT3 but not of ADAT2/A180L-ADAT3 is supported by: i) a severe decrease of the tRNAs deamination activity of the V128M and A180L complexes compared to the WT and A180V complexes *in vitro* (**Fig. 4E**) and ii) the lack of rescue of the migration phenotype with the two variants showing an impaired enzymatic activity (**Fig. 6**). Intriguingly, we also showed impaired solubility of the variants (**Fig. 4C**). However, the structural perturbations observed in the mutant complexes (V128M/L>A180L>>A180V>WT) correlate better with the defects in enzymatic activity (V128M/L; A180L> WT; A180V = WT) than the defects in solubility (A180L> A180V>>V128M/L=WT) (**Fig. 4F**). Altogether, these results suggest that, although one cannot exclude that the variants preclude the formation of the complexes *in vivo*, the structural alterations are likely one of the major determinants of the loss of deamination activity of the mutant complexes and might greatly contribute to the development of NDDs.

At the molecular level, we provided the first exhaustive quantitative analysis of the impact of ADAT3 variants on both deamination and expression of ADAT2/ADAT3 target tRNAs. Analysis of the proportion of wobble inosine (I_34_) in ANN isodecoders in cells derived from patients carrying either the p.V144M/p.V144M or p.A196V/p.A196L variants revealed striking decrease of A_34_ to I_34_ editing in about 45% of the INN isodecoders (**Fig. 5**). This extends the initial observations showing impaired deamination on few tRNA species in patient’ cells expressing the p.V144M/p.V144M variant.^16^ Of note, in contrast to ADAT2-depleted human HEK293T cells, where all the ADAT substrate tRNA families showed severe defects in deamination, ^27^ in ADAT3 patients’ cells, only the tRNA-Ala family is affected (**Supplementary** Fig.4E**-F**), suggesting cell specific effect of ADAT perturbation. Corroborating this hypothesis, it has been shown that proliferating versus differentiating cells translate the codons read by I_34_ tRNAs with different efficiency.^49^ In addition, tRNA-Ala anticodon is one of the only enriched family in the embryonic brain,^50^ suggesting further potential brain sensitivity to ADAT impairment.

Interestingly, the isodecoders of a tRNA family are not affected to the same extent (ranging from 100% I_34_ to 11% for the more severely affected isodecoders). One possible explanation for this differential effect on isodecoders within a same tRNA family is that the mutations in the mammalian ADAT2/ADAT3 complex selectively affect the binding and/or accommodation of specific tRNAs to its catalytic site. Interestingly, the structure of the *Trypanosoma brucei* ADAT2/ADAT3 complex bound to a full-length tRNA showed that the eukaryotic ADAT2/ADAT3 complex select and correctly position the tRNA substrate within the catalytic site through sequence-independent interactions.^19^ Additional biochemical analysis using chimeric tRNAs or fragments of tRNAs confirm that tRNA structural features are key for tRNA recognition by human ADAT and suggest that the mode of recognition of tRNA by ADAT2/ADAT3 complex varies between different tRNAs.^51^ A non-exclusive alternative is that I_34_ specifically stabilizes some tRNAs more than others. In such cases, some tRNAs would be found as both I_34_ and A_34_, while others would be found only as I_34_, the A_34_ species being degraded. This is supported by the facts that: i) our analysis of tRNA abundance showed that most tRNAs commonly downregulated in all three patient samples were ADAT targets (**Fig. 5**); and that ii) the extent of I_34_ loss in target tRNA molecules negatively correlate with the degree of down regulation (**Fig. 5D**). How the lack of I_34_ modification could affect stability of specific tRNAs remains unknown. While some specific structural determinants might be at play, the often very limited sequence changes between isodecoders suggest that additional mechanisms might be involved. Although none of the modifications detected by mim-tRNAseq, including m^1^A at position 58 of the tRNA-Val-AAC, Thr-AGU and Pro-AGG,^16,17^ are severely impaired in the disease context, one cannot exclude that other I_34_ dependent tRNA modifications that are known to stabilize tRNAs (*i.e.,* methylation)^52,53^ could account for the decreased steady state level of some but not other ADAT2/ADAT3 cognate tRNAs. Several evidence are in favor of a crosstalk between I_34_ and the modification of other bases. First, we previously showed that the recombinant *E. coli* TadA enzyme co-purifies exclusively with its cognate tRNA-Arg-ACG that is fully modified with I_34_ but unexpectedly also harbors an uncharacterized methylation at G_18_.^21^ Second, a recent study demonstrated that formation of m^5^C_38_ by Dnmt2 on Val-AAC depends on the pre-existing A-to-I modification at position 34.^52^ Given that methylation of C_38_ of Asp-GTC and Gly-GCC protect them against endonucleolytic cleavage,^54^ it is possible that hypomodification of Val-AAC at position 34 makes it more vulnerable to such cleavage. Therefore, full characterization of modification status of each tRNA molecules will be a strong asset to understand the differential downregulation of ADAT2/ADAT3 target tRNAs upon decreased I_34_ levels.

As the I_34_ is residing in the wobble position of the tRNA anticodon, it has profound impact upon codon-anticodon recognition and is thought to shape the proteome landscape in a codon-usage dependent manner. First, ADAT deficiency in HEK293 cells lead to impaired translation of specific proteins containing regions encoded by sequences enriched in ADAT dependent codons,^27^ in particular, proteins involved in attachment to the extracellular matrix (ECM).^27^ On the same line, inhibition of tRNA I_34_ modification in *Neurospora* specifically modulates decoding rates for ADAT-related codons with major consequence on the synthesis of ribosomal proteins.^31^ Given the key roles both the ECM and the ribosomal proteins in regulating neuronal migration,^55^ translational impairment of this subset of proteins could contribute to the neuronal migration defects observed *in vivo*. Second, computation of tRNA genes usage has suggested that the specific enrichment of tRNA-Ala-AGC anticodon in neurons drives an increased translational efficiency at Ala codons including GCC codons that are read by tRNA-Ala-I_34_GC.^50^ Given that deamination of tRNA-Ala-AGC is the most severely affected in patient cells (**Fig. 5A**), this raises the possibility that defect in decoding Ala codons contribute to neurodevelopmental phenotypes associated to mutant ADAT2/ADAT3 complex. Third, as tRNA I_34_ modification has been shown *in vitro* to be a critical determinant for recognition by the yeast isoleucyl-tRNA synthetase,^56^ defect in the I_34_ formation observed upon ADAT3 mutation might lead to accumulation of uncharged or mischarged tRNAs and subsequent translational stalling or errors, directly affecting homeostasis of proteins enriched in specific amino-acid. Collectively, these evidences converge towards translational impairment of a subset of genes rather than global translation. Therefore, the tRNAs deamination defects induced by mutant ADAT3 could impair efficiency and accuracy of translation of a subset of proteins that need to be identified to understand the sensitivity to the brain to ADAT3 loss of function.

Altogether our results demonstrate that maintaining a proper level of ADAT2/ADAT3 complex activity and subsequent level of I_34_ modification is critical for cerebral cortex development. We propose a model in which the variant alters both the solubility and the activity of the complex so that it dictates the severity of the phenotype induced by the given variant. Whether or not the threshold activity of the complexes required for proper migration or for other developmental process is the same remains to be tested. Overall, our result raised the possibility of a threshold of activity below which the tRNAs modification (I_34_) would be compatible with life but not sufficient to ensure protein demand during brain development, leading to neurodevelopmental disorders.

## Materials and Methods

### WES and patients

#### Patient 1

Patient 1 and the parents were part of the IRB (P00032816)-approved Boston Children’s Hospital (BCH) CP Sequencing Study and underwent trio-exome sequencing. Full methods regarding DNA isolation, sequencing and variant identification are described in the original paper.^33^

#### Patient 2

Genomic DNA was extracted from the peripheral blood of Patient 2 and WES was carried in Centogene labs using CentoXome® Solo. DNA was enzymatically fragmented, and target regions (including approximately 41 Mb of the human coding exome (targeting > 98% of the coding RefSeq from the human genome build GRCh37/hg19), as well as the mitochondrial genome were enriched using DNA capture probes. The generated library was sequenced on an Illumina platform to obtain at least 20x coverage depth for > 98% of the targeted bases. An in-house bioinformatics pipeline, including read alignment to GRCh37/hg19 genome assembly and revised Cambridge Reference Sequence (rCRS) of the Human Mitochondrial DNA (NC_012920), variant calling, annotation, and comprehensive variant filtering was applied. All variants with minor allele frequency (MAF) of less than 1% in gnomAD database, and disease-causing variants reported in HGMD®, in ClinVar or in CentoMD® were evaluated. All potential patterns for mode of inheritance were considered and provided family history and clinical information were used to evaluate identified variants with respect to their pathogenicity and disease causality. This study was conducted in accordance with the declaration of Helsinki 1975 for studies involving human participants and was approved by the institutional review board at faculty of medicine, Cairo University (IRB N-486-2023).

#### Patients 3 and 4

DNA from the probands was subjected to Agilent Sure-Select Human All Exon v2.0 (44Mb target) and Illumina Rapid Capture Enrichment (37Mb target) library preparation and sequenced on Illumina HiSeq 2000 or 4000 instruments. WES was performed on blood-derived DNA from two siblings. This study was approved by the Institutional Review Board at the University of California San Diego Human Research Protection Program (HRPP), San Diego, USA under protocol #140028 entitled “The Genetics of Childhood Neurological Diseases” with Dr. Gleeson as PI. Variant identification using genome analysis toolkit (GATK) workflow identified variants that were intersected with identity-by-descent blocks from Homozygosity Mapper. Variants were filtered for minor allele frequency (MAF) > 1:1000, PolyPhen-2 scores of < 0.9, or GERP score < 4.5, and runs of homozygosity were defined with Homozygosity Mapper. Potentially deleterious variants were prioritized against an in-house exome database consisting of 10,000 ethnically matched individuals, in addition to publicly available exome datasets, cumulatively numbering over 20,000 individuals. The identified mutation was confirmed by Sanger sequencing and segregation in both parents of the affected children. We prioritized predicted protein frame shift, stop codon, splice defects, and conserved nonsynonymous amino acid substitution mutations [Genomic Evolutionary Rate Profile (GERP) score > 4 or phastCons (genome conservation) score > 0.9]. We excluded variants with an allele frequency of greater than 0.2% in our internal exome database of over 2000 individuals.

#### Patients 5 and 6

Patient 5 and his family were identified and evaluated in a clinical setting, and biological samples were collected for research purposes after obtaining written informed consent according to protocols approved by Cairo University Faculty of Medicine Research Ethics Committee (N-78-2016) and Boston Children’s Hospital Institutional Review Board (05-05-076R). Trio exome sequencing and data processing for the family were done at the Genomics Platform at the Broad Institute of MIT and Harvard. An exome sequencing library was generated using the Illumina Nextera exome capture kit and sequenced using 150 bp paired-end reads to cover >80% of targets at 20X coverage and a mean target coverage of >100X. Exome sequencing data were processed using the standard pipeline with Picard. BWA^57^ was used to align reads to hg38, and SNV and insertions/deletions were called using GATK HaplotypeCaller^58^ using default filters. Variants were annotated using Variant Effect Predictor and uploaded to seqr^44^ for review. While pregnant with Patient 6 (the sister of Patient 5), the mother had prenatal diagnosis by targeted sequencing of the c.430G>A variant performed at Centogene that confirmed the homozygous variant.

#### Patient 7

Peripheral blood of Patient 7 was collected for DNA extraction after obtaining written informed consent according to protocols approved by Cairo University Faculty of Medicine Research Ethics Committee (N- 408-2023). The coding and flanking intronic regions were enriched using in solution hybridization technology and were sequenced using the Illumina HiSeq/NovaSeq system.

Copy number variations (CNV) were computed on uniquely mapping, non-duplicate, high-quality reads using an internally developed method based on sequencing coverage depth. Briefly, we used reference samples to create a model of the expected coverage that represents wet-lab biases as well as inter-sample variation. CNV calling was performed by computing the sample’s normalized coverage profile and its deviation from the expected coverage. Genomic regions are called as variants if they deviate significantly from the expected coverage.

Bioinformatics and quality control: The bioinformatics analysis began with quality control of raw sequence reads. Clean sequence reads of each sample were mapped to the human reference genome (GRCh37/hg19). Burrows-Wheeler Aligner (BWA-MEM) software was used for read alignment. Duplicate read marking, local realignment around indels, base quality score recalibration and variant calling were performed using Freebayes. Variant data was annotated with public variant databases (VefAnno, VEP). The sequencing depth and coverage for the tested sample was calculated based on the alignments. The sequencing run included in-process reference sample(s) for quality control, which passed our thresholds for sensitivity and specificity. The patient’s sample was subjected to thorough quality control measures as well, after which raw sequence reads were transformed into variants by a proprietary bioinformatics pipeline. Copy number variations (CNVS), defined as single exon or larger deletions or duplications (Del/Dups), were detected from the sequence analysis data using a proprietary bioinformatics pipeline, which processes aligned sequence reads. The difference between observed and expected sequencing depth at the targeted genomic regions was calculated and regions were divided into segments with variable DNA copy number. The expected sequencing depth was obtained by using other samples processed in the same sequence analysis as a guiding reference. Our variant classification follows the INHERITANCE Variant Classification Schemes modified from the ACMG guideline 2015. Minor modifications were made to increase reproducibility of the variant classification and improve the clinical validity of the report. Likely benign and benign variants were not reported. The pathogenicity potential of the identified variants were assessed by considering the predicted consequence, the biochemical properties of the codon change, the degree of evolutionary conservation as well as the number of reference population databases and mutation databases such as, but not limited to, the gnomAD, ClinVar, HGMD Professional and Alamut Visual. For missense variants, in silico variant prediction tools such as SIFT, PolyPhen, MutationTaster were used to assist with variant classification. In addition, the clinical relevance of any identified CNVs was evaluated by reviewing the relevant literature and databases such as Database of Genomic Variants, EXAC, gnomAD and DECIPHER. The clinical evaluation team assessed the pathogenicity of the identified variants by evaluating the information in the patient referral, reviewing the relevant literature and manually inspecting the sequencing data if needed.

#### Patient 8

Whole genome sequencing was performed after obtaining written informed consent to perform clinal and genomic studies for research purpose. Genomic DNA was enzymatically fragmented and tagged with Illumina compatible adapter sequences. The libraries were paired-end sequenced on an Illumina platform to yield an average coverage depth of ∼ 30x. A bioinformatics pipeline based on the DRAGEN pipeline from Illumina, as well as CENTOGENE’s in-house pipeline was applied. The sequencing reads were aligned to the Genome Reference Consortium Human Build 37 (GRCh37/hg19), as well as the revised Cambridge Reference Sequence (rCRS) of the Human Mitochondrial DNA (NC_012920). Sequence variants (SNVs/indels) and copy number variations (CNVs) are called using DRAGEN, Manta and in-house algorithms. Variants with a minor allele frequency (MAF) of less than 1% in gnomAD database, or disease-causing variants reported in HGMD®, in ClinVar or in CENTOGENE’s in-house Biodatabank were evaluated. Although the evaluation is focused on coding exons and flanking intronic regions, the complete gene is interrogated for candidate variants with plausible association to the phenotype. All potential modes of inheritance are considered. In addition, the provided clinical information and family history are used to evaluate identified variants with respect to their pathogenicity and disease causality. Variants were categorized into five classes (pathogenic, likely pathogenic, VUS, likely benign, and benign) according to ACMG guidelines for classification of variants in addition to ClinGen recommendations. For detection of SNVs and indels in the regions targeted for downstream analysis a sensitivity of 99.9%, a specificity of 99.9%, and an accuracy of 99.9% is achieved. CNV detection software has a sensitivity of more than 95%. CENTOGENE has established stringent quality criteria and validation processes for variants detected by NGS. Variants with low sequencing quality and/or unclear zygosity were confirmed by orthogonal methods. Consequently, a specificity of > 99.9% for all reported variants is warranted. Screening of repeat expansions is performed by the Expansion Hunter algorithm for the following genes: AR, ATN1, ATXN1, ATXN2, ATXN3, ATXN7, ATXN8OS, ATXN10, CACNA1A, CNBP, CSTB, C9ORF72, DMPK, FMR1, FXN, HTT, JPH3, NOP56, PABPN1, PHOX2B, PPP2R2B, PRNP and TBP. Screening of uniparental disomy (UPD) is performed using an in-house algorithm for Mendelian inheritance errors (MIE) to detect runs of homozygosity (ROH) for the well-known clinically relevant chromosomal regions (6q24, 7, 11p15.5, 14q32, 15q11q13, 20q13 and 20).

#### Patient 9

DNA extraction from peripheral blood of Patient 9 was performed using a commercial extraction kit (ROCHE, Germany) according to manufacturer instructions. Exome sequencing was performed using the Agilent SureSelect Target Enrichment V6 Kit, and the resulting library was sequenced on the Illumina HiSeq 2000/2500 platform. Reads were aligned to the hg19 human reference genome assembly and subjected to quality control with BWA and Sam tools. Variant calling for single nucleotide polymorphisms (SNPs) and insertions/deletions (indels) carried out using VarScan v2.3.9. Functional annotation of detected variants was conducted with Wannovar. Variants were filtered focusing on known/most relevant genes based on homozygosity, allelic frequencies in population databases, gene function impact, and predictions from various databases. Pathogenicity of detected variants was evaluated according to the American College of Medical Genetics (ACMG) guidelines. Copy number variations and mitochondrial DNA variants were not examined. Prior to blood sampling, informed consent was obtained from patient’s legal guardians according to the protocol approved by Ethics Committee of Mashhad University of Medical Sciences, Mashhad, Iran.

#### Patients 10 and 11

Peripheral blood of Patients 10 and 11 were collected for DNA extraction and establishment of lymphoblastoid cell lines. Genome sequencing and data processing were performed by the Genomics Platform at the Broad Institute of MIT and Harvard. PCR-free preparation of sample DNA (350 ng input at >2 ng/μl) is accomplished using Illumina HiSeq X Ten v2 chemistry. Libraries are sequenced to a mean target coverage of >30x. Genome sequencing data was processed through a pipeline based on Picard, using base quality score recalibration and local realignment at known indels. The BWA aligner was used for mapping reads to the human genome build 38. Single Nucleotide Variants (SNVs) and insertions/deletions (indels) are jointly called across all samples using the Genome Analysis Toolkit (GATK) HaplotypeCaller package version 4.0. Default filters were applied to SNV and indel calls using the GATK Variant Quality Score Recalibration (VQSR) approach. Annotation was performed using Variant Effect Predictor (VEP). Lastly, the variant call set was uploaded to seqr^44^ for analysis. The study protocol was approved by the Massachusetts General Brigham Institutional Review Board (Protocol #: 2016P001422) and informed consent was obtained from the participating family.

#### Patients 12 to 20

Informed consent was obtained from patient’s legal guardians according to the protocol approved by KFSHRC Ethics Committee (REC#2070023). DNA extraction from peripheral blood was performed using a commercial extraction kit (Qiagen, USA) according to manufacturer instructions. Exome sequencing was performed on genomic DNA using the Agilent SureSelect Target Enrichment workflow to capture regions of interest from a DNA fragment library. The whole exome is sequenced on the Illumina HiSeq 2500 with a minimum coverage of 30X. Reads were aligned to the human genome build UCSC hg19 genome assembly and an in-house pipeline was used to compare the proband’s sequence to the reference sequence. Coverage and quality for targeted coding exons were assessed. Analysis of Exome data was performed as described previously.^59^ Identified variants were classified according to the American College of Medical Genetics (ACMG) guidelines.

### Cloning and plasmid constructs

miRNAs against coding sequences (CDSs) for mouse *Adat3* (NM_001100606), *Adat2* (NM_025748.4), were generated using BLOCK-iT™ RNAi Designer (https://rnaidesigner.thermofisher.com/rnaiexpress/).

Sense and antisense oligos (**Supplementary Table 4**) were annealed and the resulting duplex was subcloned in pCAGGs-mir30 (Addgene plasmid # 14758)^60^ or NeuroD-miR30 vectors digested with XhoI and EcoRI. NeuroD-miR30 vector was generated by replacing the pCAGGs promoter in pCAGGSs-miR30 backbone with NeuroD promoter from the NeuroD-IRES-GFP plasmid.^61^

Wild-type (WT) mouse *Adat3 (*NCBI Reference Sequence NM_001100606) *and Adat2* (NM_025748.4 CDSs were amplified from E16.5 cortices using primers listed in **Supplementary Table 4** and cloned into pJET 1.2 blunt vector using CloneJet PCR Cloning kit. They were further subcloned into psiSTRIKE DCX- IRES-GFP (provided by J. Chelly (IGBMC, Strasbourg, France)) and pnThx^62^ *(Adat3)*, pet16B (Novagen (EMD Millipore)) (*Adat2*), NeuroD-IRES-GFP^61^ (*Adat2*), pCAGGs-IRES-GFP^63^ (*Adat2, Adat3*) vectors by restriction-ligation. For bacterial expression, the m*Adat2* gene was inserted in the pnCS vector that does not code for any fusion tag. The m*Adat3* gene was inserted in the pnEA-HT3 vector, in frame with a 5′- sequence coding for an N-terminal histidine-tag, thioredoxin and a protease 3C cleavage site.^21^ miR1*-Adat3* resistant constructs were obtained by site-directed mutagenesis using the primers indicated in **Supplementary Table 4** and subcloned into the psiSTRIKE DCX-IRES-GFP, and pCAGGs-IRES-GFP plasmids. Insensitivity of the vectors was validated by transfection of HEK239T cells together with the miRNA constructs. Mouse *Adat3* V128M, A180L and A180V variants were created from WT CDS by sequence- and ligation-independent cloning (SLIC) and subcloned into the psiSTRIKE DCX-iresGFP and pnThx ^62^ vectors. Mouse catalytically inactive Adat2 (E73A)^18^ was generated from the WT *Adat2* CDSs respectively by site-directed mutagenesis using the primers listed in **Supplementary Table 4**. All the vectors used in this study were prepared using the EndoFree plasmid purification kit (Macherey Nagel).

### Generation of Rabbit antibodies for mouse ADAT3 and ADAT2

WT mouse mADAT3 and mADAT2 full length proteins were expressed by transformation of pnThx-*Adat3*, pet16B-*Adat2* vectors^62^ into BL21 (DE3) Rosetta®(DE3) *E. coli* cells (Novagene). Bacterial cultures were grown in 2XLB media for 5-6 hours at 37°C and 200rpm. Temperature was then decreased to 22°C and recombinant protein expression was induced by addition of 0.5 mM IPTG to the LB culture media that was grown O/N at 180rpm. Next day, Cultures were harvested, resuspended in resuspension buffer (200mM NaCl, 10mM Tris pH8) and sonicated on ice. The lysate was centrifuged and the supernatant was incubated with Talon Metal Affinity Resin (Clontech) for 2h at 4°C. The resin was washed with resuspension buffer to get rid of the unbound proteins. Adat3 was eluted from the column by incubating the resin with 3C protease and Adat2 was eluted by by addition of 200mM NaCl, 10mM Tris, 250mM Imidazole (pH8) buffer. Eluted proteins were concentrated using Amicon^®^ Ultra 15ml Centrifugal Filters (Merck) and loaded on HiLoad^®^ 16/600 Superdex^®^ columns (Akta Pure, Purification system) for affinity-based protein purification. Eluted fractions containing mouse ADAT3, ADAT2 proteins were respectively pooled and dialyzed against PBS overnight using 3.5K Slide-A-Lyzer™ G2 Dialysis Cassettes (ThermoFisherScientific).

300 μg of purified recombinant WT mouse ADAT3 and ADAT2 full length proteins were used for immunization of rabbits. One month later, 40 ml of blood was drawn every week four times and the serums were collected. Rabbits were boosted with 150 μg of peptide (in a 1:1 PBS/incomplete Freund adjuvant emulsion) and killed 12 days later, under anesthesia. Antibodies were purified from serum with SulfoLink- columns coupled to the immunogens (20325, Thermo Fischer Scientific) according to manufacturer’s protocol and specificity validated by Western Blot **(Supplementary Fig. 1B)**.

### Mice

All animal studies were conducted in accordance with French regulations (EU Directive 86/609 – French Act Rural Code R 214-87 to 126) and all procedures were approved by the local ethics committee and the Research Ministry (APAFIS#15691-201806271458609 and #4220-2016022318474293). Mice were bred at the IGBMC animal facility under controlled light/dark cycles, stable temperature (19°C) and humidity (50%) condition and were provided with food and water ad libitum. Timed-pregnant WT NMRI (Janvier-labs) and CD1 (Charles River Laboratories) females were used for *in utero* electroporation of the different constructs at embryonic day 14.5 (E14.5).

### *In utero* electroporation

*In utero* electroporation (IUE) was performed as described previously.^64,65^ Briefly, CD1 pregnant females were anesthetized with isoflurane (2L/min of oxygen; 4% isoflurane in the induction phase followed by 2% isoflurane during surgery; Tem Sega). The uterine horns were exposed, and a lateral ventricle of each embryo was injected using pulled glass capillaries (Harvard apparatus, 1.0OD*0.58ID*100mmL) with Fast Green (1 µg/µl; Sigma) combined with different amounts of DNA constructs using a micro injector (Eppendorf Femto Jet). Plasmids were electroporated into the neuronal progenitors adjacent to the ventricle by 5 electric pulses (40V) for 50 ms at 950 ms intervals using electrodes (diameter 3 mm; Sonidel CUY650P3) and ECM-830 BTX square wave electroporator (VWR international). After electroporation, embryos were placed back in the abdominal cavity and the abdomen was sutured using surgical needle and thread. For E18.5 analysis, pregnant mice were sacrificed by cervical dislocation four days after surgery. For post-natal analysis, electroporated pups were sacrificed two days after birth (P2) by head sectioning. Conditions of IUE with plasmids and concentration used are summarized in **Supplementary Table 5.**

### Mouse brain fixation, cutting and immunolabelling

E18.5 and P2 animals were sacrificed by head sectioning and brains were fixed in 4% paraformaldehyde (PFA, Electron Microscopy Sciences) diluted in Phosphate buffered saline (PBS, HyClone) overnight at 4°C. WT E18.5 cryosections were prepared for immunolabeling as follows: after fixation, brains were rinsed and equilibrated in 20% sucrose in PBS overnight at 4°C, embedded in Tissue-Tek O.C.T. (Sakura), frozen on dry ice, cut coronally at the cryostat (18 µm thickness, Leica CM3050S) and maintained at -80°C until immunolabeling. For IUE analyses, vibratome section were prepared as follows: after fixation, brains were washed and embedded in a 4% low-melting agarose solution (Bio-Rad) and cut at a thickness of 60µm coronally using a vibrating-blade microtome (Leica VT1000S, Leica Microsystems). Sections were kept in PBS-azide 0.05% for short-term storage or in an antifreeze solution (30% Ethyleneglycol, 20% Glycerol, 30% DH2O, 20% PO_4_ buffer) for long-term storage. For immunolabeling cryosections and vibratome sections were permeabilized and blocked with blocking solution (5% Normal Donkey Serum (NDS, Dominic Dutscher), 0.5% Triton-X-100 in PBS) for 1h at room temperature (RT). Sections were then incubated with primary antibodies (see **Supplementary Table 6**) diluted in blocking solution overnight at 4°C and with secondary antibodies (see **Supplementary Table 6**) and DAPI (dilution 1/1000, 1mg/mL Sigma) diluted in PBS 0.1% Triton for 1h at RT. Slides were mounted using Aquapolymount mounting medium (Polysciences Inc).

### Primary neuronal culture and immunolabeling

Cortices from WT CD1 mice at E15.5 were dissected in cold PBS supplemented with BSA (3 mg/mL), MgSO4 (1 mM, Sigma), and D-glucose (30 mM, Sigma). They were enzymatically dissociated in Neurobasal medium containing papain (20 U/mL, Worthington) and DNase I (100 μg/mL, Sigma) for 20 minutes at 37°C, washed 5 minutes with Neurobasal medium containing Ovomucoide (15 mg/mL, Worthington) and manually triturated in Optimem with 20mM D-glucose. 2 x 10^5^ cells per well were plated in a 24-well plate previously coated overnight at 4°C with poly-D-lysine (1 mg/mL, Sigma). Cells were then either fixed 2h after plating or cultured in Neurobasal medium supplemented with B27, L-glutamine (2 mM) and penicillin-streptomycin (5 U/mL and 50 mg/mL, respectively) till DIV2 and fixed in 4% PFA and 4% sucrose in PBS for 15 minutes at RT. Cells were then blocked for 1 hour in 0,1% Triton X-100, 5% NDS in PBS and primary antibodies (see **Supplementary Table 6**) were added overnight at 4°C. Next day they were washed and incubated with secondary antibodies (see **Supplementary Table 6**) and DAPI (dilution 1/1000, 1mg/mL Sigma) for 1 hour at RT. Subsequently, they were mounted in Aquapolymount mounting medium (Polysciences Inc).

### Cell culture and transfections

Human embryonic kidney 293T (HEK293T) cells were cultured in Dulbecco’s modified Eagle’s medium (DMEM, GIBCO) with 10% foetal calf serum (FCS), penicillin 100 U/mL and streptomycin 100 μg/mL. Mouse neuroblastoma N2A (ATCC) cells were cultured in DMEM (GIBCO) supplemented with 5% Fetal Calf Serum (FCS) and Gentamycin 40µg/ml in a humidified atmosphere containing 5% CO2 at 37°C.

All the cells were incubated in a humidified atmosphere containing 5% CO2 at 37°C. Human lymphoblastoid cell lines (LCLs) were generated from blood samples of Patients 6 and 7 or has been previously described in^16^. LCLs were cultured in RPMI 1640 medium containing 15% fetal bovine serum, 2 mM L-alanyl-L-glutamine (GlutaMAX; Gibco), and 1% penicillin-streptomycin.

For transfection, when cells reached 40-60% confluence, they were transfected using Lipofectamine 2000 (Invitrogen) according to the manufacturer’s protocol. 48h post-transfection expression of transfected genes was assessed by RT-qPCR and western blot analysis. To assess miRNAs or shRNA knock-down efficacy and validate specificity of antibodies, HEK293T cells were transfected with 1 µg of pCAGGs-*Adat3*-IRES-GFP, pCAGGs-*Adat2*-IRES-GFP, and 3 µg of pCAGGs-miR30-scramble or pCAGGs-miR30-miRNA targeting *Adat3*, *Adat2*, respectively. To validate miRNA resistant vectors, HEK293T cells were transfected with 1 µg of either pCAGGs-*Adat3*-IRES-GFP (WT or miRNA resistant) together with 3 µg of pCAGGs- miR30-scramble or pCAGGs-miR30-miRNA targeting *Adat3*. For confirmation of expression of mutant vectors 1 µg of the respective psiSTRIKE DCX-IRES-GFP or NeuroD-IRES-GFP vectors were transfected in N2A cells.

### RNA extraction, cDNA synthesis and RT–qPCR

Total RNA from brain tissues (NMRI) or cells was extracted using TRIzol reagent (Thermo Fischer Scientific). and submitted to DNAseI treatment (TurboDNAse, ThermoFisher). cDNA samples were synthetized with SuperScript IV Reverse Transcriptase (Invitrogen) and quantitative RT-PCR (qRT-PCR) was done with amplified cDNA and SYBR Green Master Mix (Roche) together with 0.1 μM of forward and reverse primers using a Lightcycler® 480 (Roche). RT-qPCRs on WT NMRI mouse cortices from E12.5- E18.5 embryos, human LCLs and HEK293T cells transfected with different constructs were carried using the primers listed in **Supplementary Table 4**.

### Protein extraction and western blot

Proteins from mouse cortices (E12.5 to P2, NMRI), transfected cells (HEK 293T, N2A) and LCLs were extracted as follows: cells or tissue were lysed in RIPA buffer (50 mM Tris pH 8.0, 150 mM NaCl, 5 mM EDTA pH 8.0,1% Triton X-100, 0.5% sodium deoxycholate, 0.1% SDS) supplemented with EDTA-free protease inhibitors (cOmplete™, Roche) for 30 min, then cells debris were removed by high speed centrifugation at 4°C for 25 min. Protein concentration was measured by spectrophotometry using Bio-Rad Bradford protein assay reagent. Samples were denatured at 95°C for 10 min in Laemmli buffer (Bio-Rad) with 2% β-mercaptoethanol, then resolved by SDS–PAGE and transferred onto nitrocellulose membranes. Membranes were blocked in 5% milk in PBS buffer with 0.1% Tween (PBS-T) and incubated overnight at 4°C with the appropriate primary antibody in blocking solution. Membranes were washed 3 times in PBS- T, incubated at room temperature for 1 h with HRP-coupled secondary antibodies at 1:10,000 dilution in PBS-T, followed by 3 times PBS-T washes. Visualization was performed by quantitative chemiluminescence using SuperSignal West Pico PLUS Chemiluminescent Substrate (Sigma). Signal intensity was quantified using ImageQuant LAS 600 (GE Healthcare). Primary and secondary coupled HRP antibodies used for western blot are described in **Supplementary Table 6**. All immunoblot experiments consisted of at least three independent replicates.

### Small-scale expression tests of mADAT2/ADAT3

For small-scale expression tests, pnCS-mADAT2 WT and pnEA-HT3-mADAT3 WT and mutants were expressed alone or co-expressed in *Escherichia coli* BL21(DE3) cells. Transformed cells were cultivated at 37°C for 6 h in 24 deep well plates harbouring 4 mL of 2xLB Broth medium and the required antibiotics in each well. Induction was then performed overnight at 22°C by adding a final concentration of 0.7 mM of isopropyl-1-thio-β-D-galactopyranoside (IPTG) in the presence of 100 μM of Zn(SO_4_)_2_. Cells were harvested and resuspended in a buffer containing 10 mM Tris-HCl pH 8.0 and 200 mM NaCl. For total protein expression analysis, 100 μL of the resuspended cells were mixed with 10 μL of Laemmli buffer and heated for 10 minutes at 95°C prior to analysis by SDS-PAGE followed by Coomassie staining. The rest of the resuspended cells were lysed and centrifuged at 4 000 rpm for 30 m at 4°C, and the supernatants incubated with TALON Metal Affinity Resin (Clonetech) for 2h at 4°C, then centrifuged again and washed two times. For soluble protein analysis, the TALON resin was mixed with 20 μL of Laemmli buffer and heated for 10 minutes at 95°C prior to analysis by SDS-PAGE.

### Large-scale overproduction and purification of mADAT2/ADAT3

mADAT2/ADAT3 WT and mutants were produced by co-expression in *E. coli* BL21(DE3) cells. Cultures were cultivated in 2xLB Broth medium at 37°C for 6 h. Induction was then performed at 22°C overnight by adding final concentration of 0.7 mM of IPTG and 100 μM of Zn(SO_4_)_2_. Cells were harvested, resuspended and lysed in a buffer containing 10 mM Tris-HCl pH 8.0 and 200 mM NaCl and centrifuged at 17 500 rpm for 1 h at 4°C. The supernatant was incubated with TALON Metal Affinity Resin (Clonetech). To release the his-tagged complex from the TALON resin, the sample was treated with 3C protease overnight at 4°C. The next day, ion exchange chromatography was performed with a HiTrap Q HP column (GE Healthcare) using a gradient of NaCl from 50 mM to 1 M NaCl to remove bound nucleic acids. The sample was then further purified by size exclusion chromatography in 10 mM Tris HCl pH 8.0, 200 mM NaCl and 0.5 mM TCEP on a 16/600 Superdex 200 gel filtration column (GE Healthcare). The recombinant complexes were used for crystallization assays and enzymatic deamination assays.

### Protein crystallization

For crystallization, the A180V and A180L mutant ADAT complexes at 12 mg*/*ml were mixed with an equal volume of reservoir reagent and crystallized using the sitting drop vapor diffusion technique at 20°C. All crystals grew within one week. Crystals could only be obtained for the A180V mutant ADAT complex using a crystallization solution containing 0.1 M Bis-Tris-Propane pH 6.5, 20.5% PEG 3350 and 0.2 M NaBr.

### Data collection, structure determination, model building and refinement

For data collection, the crystals were frozen in liquid nitrogen after their short transfer into a cryo-protectant solution composed of their crystallization conditions added with either 20% glycerol or 20% PEG200. Data collection was performed under cryogenic conditions on beamline PXIII at the Swiss Light Source synchrotron (SLS, Switzerland) using a 1 Å wavelength. Data sets collected were processed with XDS.^66^ Structure determination was made by molecular replacement using our previous structure of the WT mADAT2/ADAT3 complex (PDB entry #7NZ8) and the structure refined by several cycles of manual building using Coot^67^ and automated refinement using Phenix.^68^ The final model was validated using tools provided in Coot and Molprobity^69^. The mADAT2/ADT3-A180V structure has been deposited in the Protein Data Bank.

### tRNA production

The mouse tRNA^Arg^(ACG) gene used was synthesized using two complementary oligonucleotides (Merk) comprising the tRNA gene sequence (underlined), a T7 RNA polymerase promoter (bold), a BstNI site (italics), and two restriction sites, HindIII and BamHI (bold underline):

5’-**AGCTTGAATTGTAATACGACTCACTATA**<u>GGGCCAGTGGCGCAATGGATAACGCGTCTGACTACGGATCAGAAGATTCTAGGTTCGACTCCTAGCTGGCTCGCC</u>*AGG***<u>G</u>**-3’ and

5’-**GATCC***CCTGG*<u>CGAGCCAGCTAGGAGTCGAACCTAGAATCTTCTGATCCGTAGTCAGACGCGTTATCCATTGCGCCACTGGCCC</u>**TATAGTGAGTCGTATTACAATTCA**-3’.

For the oligonucleotide hybridization, 4 µg of each oligonucleotide were first incubated separately at 37°C for 45 min in the presence of 4 µl of T4 DNA Ligase buffer from the T4 DNA ligase kit (Thermo Scientific^TM^ Cat.#EL0011) and 1 µl of T4 Polynucleotide Kinase (10U/µl) from the T4 Polynucleotide Kinase kit (Thermo Scientific^TM^ Cat.#EK0031) in a total volume of 25 µl. The two primer solutions were mixed, incubated at 100°C for 1 min, then at 70°C for 25 min in a water bath, and let to cool down to 20°C for two hours in the switch-off water bath. The hybridized oligos were inserted in the HindIII and BamHI-linearized pUC19 vector using the T4 DNA ligase kit (Thermo Scientific^TM^ Cat.#EL0011) according to the manufacturer’s instructions. The tRNA was synthesized from the BstNI-digested DNA by *in vitro* transcription using recombinant T7 RNA polymerase (doi.org/10.1042/BJ20121211). After transcription, the sample was treated with RQ1 RNase-Free DNase (Promega), and the RNA transcript was phenol-extracted and precipitated. Pelleted tRNA was dissolved in water and loaded on 7 M Urea-15 % acrylamide, and 1 X TBE gel. After methylene blue staining, gel slices containing the tRNA transcript were cut from the gel. The tRNA was eluted overnight at room temperature in 0.5 M ammonium acetate, 10 mM magnesium acetate, 0.1 mM EDTA and 0.1% SDS. After phenol extraction, tRNA was ethanol precipitated and finally recovered in water. The concentration was determined by absorbance measurements. The tRNA was then used for enzymatic deamination assays.

### Enzymatic deamination assays

Deamination assays were done in deamination buffer (10 mM Tris-HCl pH 8.0, 100 mM NaCl, 1 mM MgCl_2_, 2 mM dithiothreitol (DTT)) using 2 µM of tRNA transcript and 5.6, 0.56 or 0.056 µM of purified enzyme complex (tRNA:protein ratios of approximately 1:3, 1:0.3 and 1:0.003) in a final volume of 10 µL. The reaction was initiated when the purified enzyme complex was added to the reaction mixture and immediately incubated at 37°C for the indicated time (10 min). The reaction was immediately stopped by phenol-chloroform extraction. The supernatant was precipitated, and the pellet containing the tRNA transcript was dissolved in 20 µl of water. The cDNA was synthesized using the SuperScript™ IV Reverse Transcriptase (Invitrogen Cat.# 18090010) according to the manufacturer’s instructions with 2 µl of the tRNA transcript solution and 0.1 µM of gene-specific complementary primer. The tRNAs were then amplified using the GoTaq® G2 Flexi (Promega Cat.# M7801) according to the manufacturer’s instructions with 5 µl of cDNA, 2.5 mM of MgCl_2_, and 1 µM of gene-specific primers. As PCR fragments were too small for direct sequencing, they were ligated using the pGEM®-T Easy Vector System (Promega Cat.# A1360) according to the manufacturer’s instructions. The ligated PCR products were then reamplified using vector-specific and tRNA-specific primers. The resulting PCR fragments were precipitated, redissolved, and directly sequenced with the vector-specific primer. The A-to-I deamination reaction was visualized by the presence of a guanosine peak at the adenosine peak position since reverse transcriptase incorporates a cytosine instead of an inosine. The deamination rate was quantified using the peak area measurement in the Raw Data section of MacVector software (version 18.6.1). The percentage of inosine was calculated using the A and G area data sets. The forward and reverse primers specific to tRNA^Arg^ and used for RT-PCR and PCR were as follows: 5’-GGGCCAGTGGCGCAATGGA-3’ and 5’TGGCGAGCCAGCTAGGAGT-3’. The forward primer for the pGEM®-T Easy Vector was as follows: 5’-GTAAAACGACGGCCAG-3’.

### tRNA-seq library construction

LCLs derived from patients and healthy controls were lysed in lithium dodecyl sulfate (LiDS)/LET buffer (5% LiDS in 20 mM Tris, 100 mM LiCl, 2 mM EDTA, 5 mM dithiothreitol (DTT) pH 7.4 and 100 μg ml^−1^ proteinase K). The lysates were incubated at 60 °C for 10 min, pushed ten times through a 1 ml syringe with a 26 G needle and mixed by vortexing. Two volumes of cold acid phenol (pH 4.3), 1/10 volume 1-bromo-3-chloropropane and 50 µg glycogen (Thermo Fisher Scientific, AM9510) were added to the lysate and samples were mixed vigorously by vortexing, followed by centrifugation at 10,000*g* at 4 °C. The aqueous phase was transferred to a new tube and the phenol and 1-bromo-3-chloropropane extraction was repeated. RNA was then precipitated from the aqueous phase by the addition of three volumes of 100% ethanol and incubation at −20 °C for 30 min. The pellets were washed with 80% ethanol, air-dried and resuspended in RNase-free water. The RNA concentration was measured using a Nanodrop system and the samples were stored at −80 °C. The tRNASeq libraries were prepared using the mim-tRNAseq workflow ^45,46,70^. Briefly, total RNA from two biological replicates for each LCL line was mixed with synthetic *Escherichia coli* tRNA-Lys-UUU-CCA and *E. coli* tRNA-Lys-UUU-CC at a 3:1 ratio, followed by dephosphorylation with T4 PNK (NEB, M0201S) and ethanol precipitation. The RNA samples were resolved on denaturing 10% polyacrylamide, 7 M urea and 1×TBE gels. RNA of 60-100 nt was excised and eluted from the gel followed by ethanol precipitation. The gel-purified tRNA was then ligated to pre-adenylated, barcoded 3′-adaptors ^45^ in 1×T4 RNA ligase buffer, 25% PEG-8000, 20 U Superase In (Thermo Fisher Scientific, AM2696) and 1 µl T4 RNA ligase 2, truncated KQ (NEB, M0373S). The mix was incubated for 3 h at 25 °C and the ligation products were purified by size selection on a 10% polyacrylamide, 7 M urea and 1×TBE gel. Adaptor-ligated tRNA (100 ng) was annealed with 1 µl of 1.25 µM RT primer (5′ - pRNAGATCGGAAGAGCGTCGTGTAGGGAAAGAG/iSp18/GTGACTGGAGTTCAGACGTGTGCTC-3′, where iSp18 is a 18-atom hexa-ethyleneglycol spacer) at 82 °C for 2 min, followed by incubation at 25 °C for 5 min. Reverse transcription was performed with 500 nM TGIRT (InGex, TGIRT50) in 50 mM Tris–HCl pH 8.3, 75 mM KCl, 3 mM MgCl_2_, 5 mM DTT (from a freshly prepared 100 mM stock), 1.25 mM dNTPs and 20 U Superase In at 42 °C for 16 h. After reverse transcription, NaOH was added to a final concentration of 0.1 M and the RNA was hydrolyzed by incubating the samples for 5 min at 90 °C. Complementary DNA products were separated from unextended primer on a 10% polyacrylamide, 7 M urea and 1×TBE gel. Regions corresponding to cDNAs that were >10 nt longer than the RT primer were excised after SYBR Gold staining. Gel-purified and ethanol-precipitated cDNA was incubated for 3 h at 60 °C with CircLigase ssDNA ligase (Lucigen) in 1×reaction buffer supplemented with 1 mM ATP, 50 mM MgCl_2_ and 1 M betaine. Following enzyme inactivation for 10 min at 80 °C, one-fifth of the circularized cDNA was used directly for library construction PCR with a common forward (5′- AATGATACGGCGACCACCGAGATCTACACTCTTTCCCTACACGACGCT∗C-3′) and unique indexed reverse primers (5′-CAAGCAGAAGACGGCATACGAGATNNNNNNGTGACTGGAGTTCAGACGTGT∗G-3′; NNNNNN, the reverse complement of an Illumina index sequence; asterisk, phosphorothioate bond) with KAPA HiFi DNA polymerase (Roche) in 1×GC buffer with initial denaturation at 95 °C for 3 min, followed by five cycles of 98 °C for 20 s, 62 °C for 30 s and 72 °C for 30 s at a ramp rate of 3 °C s^−1^. The PCR products were purified using a DNA Clean and Concentrator 5 kit (Zymo Research), quantified with a Qubit dsDNA HS kit (Thermo Fisher Scientific, Q32851) and sequenced for 150 cycles on an Illumina NextSeq 500 platform, generating >2.5 × 10^6^ reads per library.

### Analysis of tRNASeq data

The tRNASeq data was analyzed according to the mim-tRNAseq computational workflow ^45,46,70^. Demultiplexing and 3′ sequencing adaptor removal was performed using cutadapt v3.5. Indels were disallowed (--no-indels) and both read ends were quality trimmed with a quality score of 30 (-q 30,30). As sequencing was performed with more cycles than the length of any sequenced fragment, all reads were expected to contain adaptors and only trimmed reads were retained with --trimmed-only. The reads were further trimmed to remove the two 5′-RN nucleotides introduced by circularization from the RT primer with -u 2. In both processing steps, reads <10 nt were discarded using -m 10. Analysis of tRNA expression and modification was performed with v1.3.8 of the mim-tRNAseq computational package (https://mim-trnaseq.readthedocs.io/en/latest/index.html).^46^ Briefly, the full set of 619 predicted tRNA genes for the hg38 human genome assembly were downloaded from GtRNAdb ^23^ and the 22 mitochondrially encoded human tRNA genes were fetched from mitotRNAdb ^71^. After intron removal and the addition of 5′-G (for tRNA-His) and 3′-CCA (for nuclear-encoded transcripts), a curated set of 599 nuclear-encoded tRNA sequences (excluding tRNAs with non-canonical secondary structure alignments or undetermined anticodons) and 22 mitochondrially encoded tRNA sequences was compiled as an alignment reference (--species Hsap). The reads were aligned to this reference with a cluster ID of 0.95, maximum mismatch tolerance at a number of nucleotides equal to 7.5% read length for the first alignment round and 5% read length for realignment, a deconvolution coverage ratio of 0.4 at mismatch sites to allow accurate cluster deconvolution and a minimum coverage threshold of 0.05% total reads per transcript for low coverage transcript filtering. In addition, DESeq2 was run on tRNA transcripts with single-transcript resolution by first removing those still in clusters from the counts table (evidenced by the presence of multiple transcripts in the name, separated by ‘/’) and repeating DESeq2 analysis on these. Isotype counts, generated by aggregating anticodon counts for the same tRNA isotype were also generated, and DESeq2 was additionally run on this count data.

Inosine 34 proportions were obtained from the mismatch analysis integrated in the mimtRNA-seq pipeline. For each sample, mismatch values for the canonical position 34 matching the nucleotide G were obtained and averaged across replicates as follows. For the anticodon-level analysis, the mean across all isodecoders of the same isotype weighted by their coverage was calculated and then averaged across replicates. For the isodecoder-level analysis, the mean across replicates was weighted by their total coverage. For all other modifications, given the absent of a clear mis-read designed nucleotide, the misincorporation rate at the 7 canonical positions resolved from mimtRNAseq for the ADAT-target genes (position 9 = m^1^G or m^1^A, position 20 = acp^3^U, position 26 = m^2,2^G, position 32 = m^3^C, position 37 = yW or m^1^I, position 58 = m^1^A) was calculated for each condition as for I34. tRNAs never modified in the controls in such positions (misincorporation proportion < 1%) were removed.

### Image acquisition and analysis

Images for primary neuronal culture and expression pattern analyses were acquired using a TCS SP8 UV (Leica microsystems) using a 63x OIL HC PL APO CS2 and 20x IMM, HC PL APO CS2 objectives respectively and images for neuronal migration and expression pattern analyses were acquired using a TCS SP8 X (Leica microsystems) confocal microscope using a 20x DRY HC PL APO CS2 objective. For all experiments, a Z-stack of 1,50 μm was acquired. The image size was 512x512 for neuronal migration analysis and 1024x1024 for primary neuronal culture and expression analysis. Image analysis was done using ImageJ software (NIH). Cell counting was performed in 2 to 4 different brain sections of at least 3 different embryos or pups per condition. Only similarly electroporated regions were considered for further analysis. Cortical areas (upper cortical plate, lower cortical plate, intermediate zone, subventricular zone/ventricular zone) were delimited based on cell density (nuclei count with DAPI staining) using equivalent sized boxes. Number of GFP-positive cells was determined in each cortical area to establish the percentage of positive cells. All the experiments were done in at least three independent replicates.

### Statistics

All statistics analyses were performed using GraphPad Prism 6 (GraphPad) and are represented as mean +/- S.E.M. The level of significance was set at P < 0.05 in all the statistical tests. All statistical tests used and n size numbers are shown in the figure legends and statistical details are reported in **Supplementary Table 7**. Correlation analysis was performed with R v. 4.4.0 (within R studio) and its package stats. Analysis of IUE experiments and neuroanatomical characterization of mouse brains were performed blinded. Normality was checked using Shapiro-Wilk or KS normality test depending on the sample size, when the data was big enough. In case normality was violated, non-parametric tests were used.

## Data Availability

All other relevant data included in the article are available from the authors upon request. High-throughput sequencing data has been deposited in the Gene Expression Omnibus Database (GSE278536).

## Acknowledgments

This work was supported by grants from INSERM (ATIP-Avenir program, J.D.G.), the French state funds through the Agence Nationale de la Recherche (ANR-21-CE12-0026 to J.D.G., C.R. and L.D.; ANR-DFG ANR-19-CE16-0023 to J.D.G. and D.D.N.; ANR-10-IDEX-0002-02 and ANR-10-LABX-0030-INRT to J.D.G. and C.R.), ERC CoG 2021 TransNeuroFate (J.G.), SFRI-STRAT’US project [ANR 20-SFRI-0012]; and EUR IMCBio [ANR-17-EURE-0023]), INSERM/CNRS (J.D.G., C.R, and L.D.) and University of Strasbourg (J.D.G., C.R., and L.D.). This work benefited from the French Infrastructure for Integrated Structural Biology (FRISBI; ANR-10-INBS-0005-01). D.D.N. is funded by the Max Planck Society, the European Research Council under the European Union’s Horizon 2020 Research and Innovation Programme (ERC Starting Grant 803825-TransTempoFold) and the EMBO Young Investigator Program (YIP 4833). E.B. was supported by INSERM, Fondation Jerôme Lejeune and ANR. R.L. is supported by Ministère de l’Enseignement Supérieur de la Recherche et de l’Innovation. E.Br is supported by Fondation pour la recherche médicale. P.T. is research assistant at the University of Strasbourg. T.S-G. is a CNRS research engineer. L.M. Was supported by ANR. C.R. is a CNRS investigator. J.D.G. is an INSERM investigator. Sequencing and analysis were provided by the Broad Institute of MIT and Harvard Center for Mendelian Genomics (Patients 5, 6, 7, 10 and 11) and was funded by the National Human Genome Research Institute (NHGRI), the National Eye Institute, and the National Heart, Lung and Blood Institute grant UM1HG008900 and in part by NHGRI grants R01HG009141 and U01HG011755. CAW is supported by a grant from the NINDS (R01 NS035129) and is an Investigator of the Howard Hughes Medical Institute. We acknowledge the Imaging Center of IGBMC (https://ici.igbmc.fr/) part of the IBiSA labeled PIQ-QuESt (https://piq.unistra.fr) and member of the national infrastructure France-Bioimaging (https://france-bioimaging.org) supported by the French National Research Agency (ANR-10-INBS-04). We thank members of the SLS synchrotron for the use of their beamline facility and help during data collection. We thank Edouard Troesch for help with cloning and Katrin Strasser for assistance with tRNA sequencing. High-throughput sequencing was partly performed at the NGS Facility in the Department of Totipotency at the MPI of Biochemistry. We are grateful to the staff of the molecular biology service (in particular, Thierry Lerouge and Paola Rossolillo), of the mouse facilities of the Institut Clinique de la souris (ICS) and IGBMC, and of the IGBMC cell culture service for their involvement in the project. We also thank Doulaye Dembele from the IGBMC GenomEast sequencing platform (part of France Genomic infrastructure) for his help with statistical analysis. We are also grateful to members of J.D.G.’s laboratory for discussion and technical assistance.

## Author contributions

J.D.P.R., R.L. and E.B. conceived and designed the experiments, performed the experiments, performed statistical analysis and analyzed the data related to cellular, and functional studies in mice. P.T. conceived and performed *in utero* electroporation, collected the mouse samples, processed the tissues and did immunostainings, took care of mouse colonies, coordinated *in vivo* experiments and provided technical assistance. E.Br. performed analysis of sequencing data and helped preparing the figures. H.R.V., M.V.G. and E.R-M. purified recombinant complexes and performed co-expression assays. N.S. helped with Western-blotting. H.R.V. and C.R. performed all the structural analysis. C.R. led the structural work and contributed to the writing of the manuscript. L.M. produced the tRNA, performed *in vitro* deamination assays, and analyzed the results. T.S-G. and L.D. designed and planned the in vitro deamination assays and analyzed the results. D.N. and T.B. performed and analyzed the mim-tRNAseq. G.V., E.M.E, A.K.L., M.O’L., M.C., N.M.O., D.G., A.A., M.K., D.S.A-A., W.E., N.A., A.O’D.L., J.E.N, J.G.G., C.A.W., N.E., L.S., S.S. followed-up the patients and families, provided the clinical and imaging data and/or contributed to the generation of whole-exome sequencing, bioinformatics tools and analysis of sequencing data. E.B. and J.D.G. conceived, coordinated and supervised the study and wrote the manuscript with contributions from all other authors.

## References

1. Cantara WA, Crain PF, Rozenski J, et al. The RNA Modification Database, RNAMDB: 2011 update. Nucleic Acids Res. Jan 2011;39(Database issue):D195–201. doi:10.1093/nar/gkq1028

2. Cappannini A, Ray A, Purta E, et al. MODOMICS: a database of RNA modifications and related information. 2023 update. Nucleic Acids Res. Jan 5 2024;52(D1):D239–D244. doi:10.1093/nar/gkad1083

3. de Crecy-Lagard V, Boccaletto P, Mangleburg CG, et al. Matching tRNA modifications in humans to their known and predicted enzymes. Nucleic Acids Res. Mar 18 2019;47(5):2143–2159. doi:10.1093/nar/gkz011

4. Phizicky EM, Alfonzo JD. Do all modifications benefit all tRNAs? FEBS Lett. Jan 21 2010;584(2):265–71. doi:10.1016/j.febslet.2009.11.049

5. Agris PF, Vendeix FA, Graham WD. tRNA’s wobble decoding of the genome: 40 years of modification. J Mol Biol. Feb 9 2007;366(1):1–13. doi:10.1016/j.jmb.2006.11.046

6. Suzuki T. The expanding world of tRNA modifications and their disease relevance. Nat Rev Mol Cell Biol. Jun 2021;22(6):375–392. doi:10.1038/s41580-021-00342-0

7. Arrondel C, Missoury S, Snoek R, et al. Defects in t(6)A tRNA modification due to GON7 and YRDC mutations lead to Galloway-Mowat syndrome. Nat Commun. Sep 3 2019;10(1):3967. doi:10.1038/s41467-019-11951-x

8. Blanco S, Dietmann S, Flores JV, et al. Aberrant methylation of tRNAs links cellular stress to neuro- developmental disorders. EMBO J. Sep 17 2014;33(18):2020–39. doi:10.15252/embj.201489282

9. Braun DA, Rao J, Mollet G, et al. Mutations in KEOPS-complex genes cause nephrotic syndrome with primary microcephaly. Nat Genet. Oct 2017;49(10):1529–1538. doi:10.1038/ng.3933

10. de Brouwer APM, Abou Jamra R, Kortel N, et al. Variants in PUS7 Cause Intellectual Disability with Speech Delay, Microcephaly, Short Stature, and Aggressive Behavior. Am J Hum Genet. Dec 6 2018;103(6):1045–1052. doi:10.1016/j.ajhg.2018.10.026

11. Kojic M, Abbassi NEH, Lin TY, et al. A novel ELP1 mutation impairs the function of the Elongator complex and causes a severe neurodevelopmental phenotype. J Hum Genet. Jul 2023;68(7):445–453. doi:10.1038/s10038-023-01135-3

12. Kojic M, Gaik M, Kiska B, et al. Elongator mutation in mice induces neurodegeneration and ataxia-like behavior. Nat Commun. Aug 10 2018;9(1):3195. doi:10.1038/s41467-018-05765-6

13. Kojic M, Gawda T, Gaik M, et al. Elp2 mutations perturb the epitranscriptome and lead to a complex neurodevelopmental phenotype. Nat Commun. May 11 2021;12(1):2678. doi:10.1038/s41467-021-22888-5

14. Lentini JM, Alsaif HS, Faqeih E, Alkuraya FS, Fu D. DALRD3 encodes a protein mutated in epileptic encephalopathy that targets arginine tRNAs for 3-methylcytosine modification. Nat Commun. May 19 2020;11(1):2510. doi:10.1038/s41467-020-16321-6

15. Nagayoshi Y, Chujo T, Hirata S, et al. Loss of Ftsj1 perturbs codon-specific translation efficiency in the brain and is associated with X-linked intellectual disability. Sci Adv. Mar 2021;7(13)doi:10.1126/sciadv.abf3072

16. Ramos J, Han L, Li Y, et al. Formation of tRNA Wobble Inosine in Humans Is Disrupted by a Millennia-Old Mutation Causing Intellectual Disability. Mol Cell Biol. Oct 1 2019;39(19)doi:10.1128/MCB.00203-19

17. Ramos J, Proven M, Halvardson J, et al. Identification and rescue of a tRNA wobble inosine deficiency causing intellectual disability disorder. RNA. Aug 6 2020;doi:10.1261/rna.076380.120

18. Gerber AP, Keller W. An adenosine deaminase that generates inosine at the wobble position of tRNAs. Science. Nov 5 1999;286(5442):1146-9.

19. Dolce LG, Zimmer AA, Tengo L, et al. Structural basis for sequence-independent substrate selection by eukaryotic wobble base tRNA deaminase ADAT2/3. Nat Commun. Nov 8 2022;13(1):6737. doi:10.1038/s41467-022-34441-z

20. Liu X, Chen R, Sun Y, et al. Crystal structure of the yeast heterodimeric ADAT2/3 deaminase. BMC Biol. Dec 3 2020;18(1):189. doi:10.1186/s12915-020-00920-2

21. Ramos-Morales E, Bayam E, Del-Pozo-Rodriguez J, et al. The structure of the mouse ADAT2/ADAT3 complex reveals the molecular basis for mammalian tRNA wobble adenosine-to-inosine deamination. Nucleic Acids Res. Jun 21 2021;49(11):6529–6548. doi:10.1093/nar/gkab436

22. Crick FH. Codon--anticodon pairing: the wobble hypothesis. J Mol Biol. Aug 1966;19(2):548–55. doi:10.1016/s0022-2836(66)80022-0

23. Chan PP, Lowe TM. GtRNAdb: a database of transfer RNA genes detected in genomic sequence. Nucleic Acids Res. Jan 2009;37(Database issue):D93-7. doi:10.1093/nar/gkn787

24. Grosjean H, de Crecy-Lagard V, Marck C. Deciphering synonymous codons in the three domains of life: co-evolution with specific tRNA modification enzymes. FEBS Lett. Jan 21 2010;584(2):252–64. doi:10.1016/j.febslet.2009.11.052

25. Zhou W, Karcher D, Bock R. Identification of enzymes for adenosine-to-inosine editing and discovery of cytidine-to-uridine editing in nucleus-encoded transfer RNAs of Arabidopsis. Plant Physiol. Dec 2014;166(4):1985–97. doi:10.1104/pp.114.250498

26. Wolf J, Gerber AP, Keller W. tadA, an essential tRNA-specific adenosine deaminase from Escherichia coli. EMBO J. Jul 15 2002;21(14):3841–51. doi:10.1093/emboj/cdf362

27. Torres AG, Rodriguez-Escriba M, Marcet-Houben M, et al. Human tRNAs with inosine 34 are essential to efficiently translate eukarya-specific low-complexity proteins. Nucleic Acids Res. Jul 9 2021;49(12):7011–7034. doi:10.1093/nar/gkab461

28. Torres AG, Pineyro D, Rodriguez-Escriba M, et al. Inosine modifications in human tRNAs are incorporated at the precursor tRNA level. Nucleic Acids Res. May 26 2015;43(10):5145–57. doi:10.1093/nar/gkv277

29. Rubio MA, Pastar I, Gaston KW, et al. An adenosine-to-inosine tRNA-editing enzyme that can perform C-to-U deamination of DNA. Proc Natl Acad Sci U S A. May 8 2007;104(19):7821–6. doi:10.1073/pnas.0702394104

30. Tsutsumi S, Sugiura R, Ma Y, et al. Wobble inosine tRNA modification is essential to cell cycle progression in G(1)/S and G(2)/M transitions in fission yeast. J Biol Chem. Nov 16 2007;282(46):33459–65. doi:10.1074/jbc.M706869200

31. Lyu X, Yang Q, Li L, et al. Adaptation of codon usage to tRNA I34 modification controls translation kinetics and proteome landscape. PLoS Genet. Jun 2020;16(6):e1008836. doi:10.1371/journal.pgen.1008836

32. Alazami AM, Hijazi H, Al-Dosari MS, et al. Mutation in ADAT3, encoding adenosine deaminase acting on transfer RNA, causes intellectual disability and strabismus. J Med Genet. Jul 2013;50(7):425–30. doi:10.1136/jmedgenet-2012-101378

33. Chopra M, Gable DL, Love-Nichols J, et al. Mendelian etiologies identified with whole exome sequencing in cerebral palsy. Ann Clin Transl Neurol. Feb 2022;9(2):193–205. doi:10.1002/acn3.51506

34. El-Hattab AW, Saleh MA, Hashem A, et al. ADAT3-related intellectual disability: Further delineation of the phenotype. Am J Med Genet A. Feb 3 2016;doi:10.1002/ajmg.a.37578

35. Hengel H, Buchert R, Sturm M, et al. First-line exome sequencing in Palestinian and Israeli Arabs with neurological disorders is efficient and facilitates disease gene discovery. Eur J Hum Genet. Mar 25 2020;doi:10.1038/s41431-020-0609-9

36. Sharkia R, Zalan A, Jabareen-Masri A, Zahalka H, Mahajnah M. A new case confirming and expanding the phenotype spectrum of ADAT3-related intellectual disability syndrome. European Journal of Medical Genetics. 2018;doi:10.1016/j.ejmg.2018.10.001

37. Yahia A, Ayed IB, Hamed AA, et al. Genetic diagnosis in Sudanese and Tunisian families with syndromic intellectual disability through exome sequencing. Ann Hum Genet. Jul 2022;86(4):181–194. doi:10.1111/ahg.12460

38. AlAbdi L, Maddirevula S, Shamseldin HE, et al. Diagnostic implications of pitfalls in causal variant identification based on 4577 molecularly characterized families. Nat Commun. Aug 29 2023;14(1):5269. doi:10.1038/s41467-023-40909-3

39. Salehi Chaleshtori AR, Miyake N, Ahmadvand M, Bashti O, Matsumoto N, Noruzinia M. A novel 8- bp duplication in ADAT3 causes mild intellectual disability. Hum Genome Var. 2018;5:7. doi:10.1038/s41439-018-0007-9

40. Thomas E, Lewis AM, Yang Y, Chanprasert S, Potocki L, Scott DA. Novel Missense Variants in ADAT3 as a Cause of Syndromic Intellectual Disability. J Pediatr Genet. Dec 2019;8(4):244–251. doi:10.1055/s-0039-1693151

41. Spears JL, Rubio MA, Gaston KW, et al. A single zinc ion is sufficient for an active Trypanosoma brucei tRNA editing deaminase. J Biol Chem. Jun 10 2011;286(23):20366–74. doi:10.1074/jbc.M111.243568

42. Sobreira N, Schiettecatte F, Valle D, Hamosh A. GeneMatcher: a matching tool for connecting investigators with an interest in the same gene. Hum Mutat. Oct 2015;36(10):928–30. doi:10.1002/humu.22844

43. Philippakis AA, Azzariti DR, Beltran S, et al. The Matchmaker Exchange: a platform for rare disease gene discovery. Hum Mutat. Oct 2015;36(10):915–21. doi:10.1002/humu.22858

44. Pais LS, Snow H, Weisburd B, et al. seqr: A web-based analysis and collaboration tool for rare disease genomics. Hum Mutat. Jun 2022;43(6):698–707. doi:10.1002/humu.24366

45. Behrens A, Nedialkova DD. Experimental and computational workflow for the analysis of tRNA pools from eukaryotic cells by mim-tRNAseq. STAR Protoc. Sep 16 2022;3(3):101579. doi:10.1016/j.xpro.2022.101579

46. Behrens A, Rodschinka G, Nedialkova DD. High-resolution quantitative profiling of tRNA abundance and modification status in eukaryotes by mim-tRNAseq. Mol Cell. Apr 15 2021;81(8):1802–1815.e7. doi:10.1016/j.molcel.2021.01.028

47. Elias Y, Huang RH. Biochemical and structural studies of A-to-I editing by tRNA:A34 deaminases at the wobble position of transfer RNA. Biochemistry. Sep 13 2005;44(36):12057–65. doi:10.1021/bi050499f

48. Behrens A, Rodschinka G, Nedialkova DD. High-resolution quantitative profiling of tRNA abundance and modification status in eukaryotes by mim-tRNAseq. Mol Cell. Feb 5 2021;doi:10.1016/j.molcel.2021.01.028

49. Bornelov S, Selmi T, Flad S, Dietmann S, Frye M. Codon usage optimization in pluripotent embryonic stem cells. Genome Biol. Jun 7 2019;20(1):119. doi:10.1186/s13059-019-1726-z

50. Gao W, Gallardo-Dodd CJ, Kutter C. Cell type-specific analysis by single-cell profiling identifies a stable mammalian tRNA-mRNA interface and increased translation efficiency in neurons. Genome Res. Dec 2 2021;doi:10.1101/gr.275944.121

51. Roura Frigole H, Camacho N, Castellvi Coma M, et al. tRNA deamination by ADAT requires substrate-specific recognition mechanisms and can be inhibited by tRFs. RNA. May 2019;25(5):607–619. doi:10.1261/rna.068189.118

52. Huang ZX, Li J, Xiong QP, Li H, Wang ED, Liu RJ. Position 34 of tRNA is a discriminative element for m5C38 modification by human DNMT2. Nucleic Acids Res. Dec 16 2021;49(22):13045–13061. doi:10.1093/nar/gkab1148

53. Tuorto F, Liebers R, Musch T, et al. RNA cytosine methylation by Dnmt2 and NSun2 promotes tRNA stability and protein synthesis. Nat Struct Mol Biol. Sep 2012;19(9):900–5. doi:10.1038/nsmb.2357

54. Tuorto F, Herbst F, Alerasool N, et al. The tRNA methyltransferase Dnmt2 is required for&#xa0;accurate polypeptide synthesis during&#xa0;haematopoiesis. The EMBO Journal. 2015/09/14 2015;34(18):2350-2362-2362. 10.15252/embj.201591382

55. 55. Amin S, Borrell V. The Extracellular Matrix in the Evolution of Cortical Development and Folding. Front Cell Dev Biol. 2020;8:604448. doi:10.3389/fcell.2020.604448

56. Senger B, Auxilien S, Englisch U, Cramer F, Fasiolo F. The modified wobble base inosine in yeast tRNAIle is a positive determinant for aminoacylation by isoleucyl-tRNA synthetase. Biochemistry. Jul 8 1997;36(27):8269–75. doi:10.1021/bi970206l

57. Li H, Durbin R. Fast and accurate short read alignment with Burrows-Wheeler transform. Bioinformatics. Jul 15 2009;25(14):1754–60. doi:10.1093/bioinformatics/btp324

58. McKenna A, Hanna M, Banks E, et al. The Genome Analysis Toolkit: a MapReduce framework for analyzing next-generation DNA sequencing data. Genome Res. Sep 2010;20(9):1297–303. doi:10.1101/gr.107524.110

59. Monies D, Abouelhoda M, Assoum M, et al. Lessons Learned from Large-Scale, First-Tier Clinical Exome Sequencing in a Highly Consanguineous Population. Am J Hum Genet. Oct 3 2019;105(4):879. doi:10.1016/j.ajhg.2019.09.019

60. Matsuda T, Cepko CL. Controlled expression of transgenes introduced by in vivo electroporation. Proc Natl Acad Sci U S A. Jan 16 2007;104(3):1027–32. doi:10.1073/pnas.0610155104

61. Hand R, Polleux F. Neurogenin2 regulates the initial axon guidance of cortical pyramidal neurons projecting medially to the corpus callosum. Neural Dev. Aug 24 2011;6:30. doi:10.1186/1749-8104-6-30

62. Diebold ML, Fribourg S, Koch M, Metzger T, Romier C. Deciphering correct strategies for multiprotein complex assembly by co-expression: application to complexes as large as the histone octamer. J Struct Biol. Aug 2011;175(2):178–88. doi:10.1016/j.jsb.2011.02.001

63. Nguyen L, Besson A, Heng JI, et al. p27kip1 independently promotes neuronal differentiation and migration in the cerebral cortex. Genes Dev. Jun 1 2006;20(11):1511–24. doi:10.1101/gad.377106

64. Godin JD, Thomas N, Laguesse S, et al. p27(Kip1) Is a Microtubule-Associated Protein that Promotes Microtubule Polymerization during Neuron Migration. Dev Cell. Oct 16 2012;23(4):729–44. doi:10.1016/j.devcel.2012.08.006

65. Laguesse S, Creppe C, Nedialkova DD, et al. A Dynamic Unfolded Protein Response Contributes to the Control of Cortical Neurogenesis. Dev Cell. Dec 7 2015;35(5):553–67. doi:10.1016/j.devcel.2015.11.005

66. Kabsch W. XDS. Acta crystallographica Section D, Biological crystallography. Feb 2010;66(Pt 2):125–32. doi:10.1107/s0907444909047337

67. Emsley P, Lohkamp B, Scott WG, Cowtan K. Features and development of Coot. Acta crystallographica Section D, Biological crystallography. Apr 2010;66(Pt 4):486–501. doi:10.1107/s0907444910007493

68. Liebschner D, Afonine PV, Baker ML, et al. Macromolecular structure determination using X-rays, neutrons and electrons: recent developments in Phenix. *Acta crystallographica Section D*, Structural biology. Oct 1 2019;75(Pt 10):861–877. doi:10.1107/s2059798319011471

69. Williams CJ, Headd JJ, Moriarty NW, et al. MolProbity: More and better reference data for improved all-atom structure validation. Protein science : a publication of the Protein Society. Jan 2018;27(1):293–315. doi:10.1002/pro.3330

70. Gao L, Behrens A, Rodschinka G, et al. Selective gene expression maintains human tRNA anticodon pools during differentiation. Nature Cell Biology. 2024/01/08 2024;doi:10.1038/s41556-023-01317-3

71. Jühling F, Mörl M, Hartmann RK, Sprinzl M, Stadler PF, Pütz J. tRNAdb 2009: compilation of tRNA sequences and tRNA genes. Nucleic Acids Res. Jan 2009;37(Database issue):D159-62. doi:10.1093/nar/gkn772

